# A genome-wide association study of anti-Müllerian hormone (AMH) levels in Samoan women

**DOI:** 10.1101/2024.12.05.24318457

**Authors:** Z Erdogan-Yildirim, JC Carlson, M Krishnan, JZ Zhang, G Lambert-Messerlian, T Naseri, S Viali, NL Hawley, ST McGarvey, DE Weeks, RL Minster

**Author notes:** First and corresponding author. Current affiliations: Department of Biobehavioral Health, College of Health and Human Development, Pennsylvania State University, State College, PA, USA, and Department of Epidemiology, University of North Carolina at Chapel Hill, NC, USA.

## Abstract

**Study question:** Can a genome-wide association study (GWAS) and transcriptome-wide association study (TWAS) help identify genetic variation or genes associated with circulating anti-Müllerian hormone (AMH) levels in Samoan women?

**Summary answer:** We identified eleven genome-wide suggestive loci (strongest association signal in *ARID3A* 19-946163-G-C [*p* = 2.32 × 10⁻⁷]) and seven transcriptome-wide significant genes (*GINS2, SENP3, USP7, TUSC3, MAFA, METTL4, NDFIP1* [all with a *p* < 2.50 × 10⁻⁶]) associated with circulating AMH levels in Samoan women.

**What is known already:** Three prior GWASs of AMH levels identified eight loci in premenopausal women of European ancestry *(AMH, MCM8, TEX41*, *CHECK2, CDCA7*, *EIF4EBP1, BMP4* and an uncharacterized non-coding RNA gene *CTB-99A3.1*), among which the *MCM8* locus was shared among all three studies.

**Study design, size, duration:** We included a sample of 1,185 women from two independently recruited samples: a family study (*n* = 212; [age: 18 to 40 years]) recruited in 2002–03 from Samoa and American Samoa; and the Soifua Manuia Study (*n* = 973; age: 25 to 51 years), a crosssectional population-based study recruited in 2010 from Samoa.

**Participants/materials, setting, methods:** Serum AMH levels were measured using enzyme linked immunosorbent assays (ELISA). We performed GWASs in the two participant samples using a Cox mixed-effects model to account for AMH levels below detectable limits and adjusted for centered age, centered age², polity, and kinship via kinship matrix. The summary statistics were then meta-analyzed using a fixed-effect model. We annotated the variants with *p <* 1 × 10⁻⁵ and calculated posterior probability of causality for prioritization. We further annotated variants using FUMA and performed colocalization and transcriptome-wide association analysis. We also assessed whether any previously reported loci were replicated in our GWAS.

**Main results and the role of chance:** We identified eleven novel genome-wide suggestive loci (*p* < 1 × 10⁻⁵) associated with AMH levels and replicated *EIF4EBP1,* a previously reported AMH locus, in the GWAS. The lead variant in *ARID3A*, 19-946163-G-C is in high linkage disequilibrium (*r* ² = 0.79) with the known age-at-menopause variant 19-950694-G-A. Nearby *KISS1R* is a biologically plausibility causal gene in the region; kisspeptin regulates ovarian follicle development and has been linked to AMH levels. Further investigation of the *ARID3A* locus is warranted.

**Limitations, reasons for caution:** The main limitations of our study are the small sample size for a GWAS and the use of the transcription model trained on mostly European samples from the Genotype Tissue Expression (GTEx) project, which may have led to reduced power to detect genotype-expression associations. Our findings need to be validated in larger Polynesian cohorts.

**Wider implications of the findings:** In addition to replicating one of the eight previously discovered AMH loci, we identified new suggestive associations. It is known that the inclusion of founder populations aids in the discovery of novel loci. These findings could enhance our understanding of AMH and AMH-related reproductive phenotypes (ovarian reserve, age at menopause, premature ovarian failure, and polycystic ovary syndrome) and help build a screening approach for women at risk for these phenotypes using genetically predicted AMH levels.

**Study funding/competing interest(s):** This work was funded by NIH grants R01-HL093093 (PI: S.T.M.), R01-HL133040 (PI: R.L.M.), and T90-DE030853 (PI: C.S. Sfeir). Molecular data for the Trans-Omics in Precision Medicine (TOPMed) Program was supported by the National Heart, Lung and Blood Institute (NHLBI). The content is solely the responsibility of the authors and does not represent the official views of the National Institutes of Health.

## Introduction

Anti-Müllerian hormone (AMH) has an important role in ovarian biology and female reproductive health.[1] AMH is produced in women after birth and is exclusively secreted by the granulosa cells of developing ovarian follicles until the antral stage is reached [2–4]. It functions to promote the development of a dominant follicle while suppressing non-dominant follicles. Not surprisingly, serum AMH concentrations highly correlate with the number of ovarian follicles [5–8], making it one of the best-known biomarkers of ovarian reserve (defined as the capacity of the ovary to provide egg cells that are capable of fertilization resulting in a healthy and successful pregnancy) and a predictor of the oocyte outcomes of *in vitro* fertilization [9,10]. Serum AMH concentrations reflect the trajectory of female reproductive life span, peaking around age 25 and then gradually decreasing until becoming undetectable before menopause [9,11]. Hence, AMH also has utility in predicting time-to-menopause [5–8,12–14].

In recent years, researchers have focused on the clinical application of AMH as a surrogate marker to evaluate polycystic ovaries and to diagnose polycystic ovary syndrome (PCOS), a common disorder affecting fertility and metabolic health of women [11,13,15]. Studies have consistently shown that AMH levels are higher in all PCOS subtypes compared to normo-ovulatory women and women with polycystic ovarian morphology alone [16] and that AMH levels correlate with PCOS subtype and severity [17,18]. Hence, AMH could provide a non-invasive alternative to transvaginal ultrasound for antral follicle counts, particularly when the latter is not available or not feasible due to cost and or acceptability (i.e. due to cultural and psycho-social reasons, especially for adolescents) [19].

Our research group has been focused on the health of Samoans, a founder population, for more than thirty years. We have conducted several epidemiological studies to describe the influences of adiposity on Samoan women’s reproductive health, specifically describing menstrual irregularity, hyperandrogenemia, and estimating prevalence of PCOS [20–22]. This is especially important given the high and rising levels of adiposity among Samoan women that are characteristic of Pacific Islanders more broadly [23–27]. Recently, two distinct PCOS subtypes (metabolic vs. reproductive) with distinct genetic architecture have been described in individuals of European ancestry [28], and it was found that PCOS susceptibility loci differ between lean and overweight/obese cases [29]. We do not have diagnoses of PCOS in Samoan women, and so we cannot directly examine the genetic determinants of PCOS in this study. However, it could be fruitful to examine related phenotypes to begin to understand genetic determinants of reproductive health in this population.

There have been several studies of the genetic variation underlying AMH levels [30–32]. In addition to polymorphisms within the *AMH* gene itself [32–34], GWASs have mapped seven genes in women of European ancestry, of which four are implicated in cell cycle regulation (*MCM8* [31,32,34], *TEX41* [32,34], *CHECK2* [34], and *CDCA7* [32]). The other three genes are *EIF4EBP1* [34], *BMP4* [34], and an uncharacterized non-coding RNA gene (*CTB-99A3.1* [32]). The established loci explain about 13% to 15% of the single nucleotide variant (SNV)-based heritability [32,34]. A major limitation of the existing studies, however, is that the information on the genetic underpinnings of AMH have been derived solely from women of European ancestry [35,36]. More research is needed in diverse populations not only to enhance our understanding of the underlying biology but also to ensure access to adequate health care and effective treatment for these communities. Importantly, the inclusion of founder populations in genetic research is important since the reduced allelic heterogeneity in these groups can be advantageous to discover novel loci via genome-wide association studies [23–27].

In this study, we aimed to identify genetic determinants of circulating AMH levels via genome-wide and transcriptome-wide analyses in Samoan women.

## Materials and methods

### Study subjects

Two independent study samples comprised of a total of 1,185 Samoan women were selected for a GWAS to assess the genetic variation associated with circulating serum AMH levels. The first sample of 212 women aged ≥ 18 years and < 40 years was drawn from a 2002–2003 family study of genetic linkage analysis of cardiometabolic traits (for sample flowchart see Supplementary Figure S1) [22,37,38]. The age range was limited to reproductive-aged women in the parent study [22] to avoid potential effects of perimenopause. Participants were recruited from villages across ‘Upolu and Savai‘i, the two largest islands of Samoa, and Tutuila, the largest island of American Samoa.

The second sample of 973 Samoan women aged ≥ 25 to ≤ 50 years was drawn from a 2010 cross-sectional population-based study (Soifua Manuia [in Samoan: “Good Health”] Study) of obesity and cardiometabolic health [21,24,39] (for sample see Supplementary Figure S1). Participants were recruited from thirty-three villages across ‘Upolu and Savai‘i. All participants completed a questionnaire surveying their health history and lifestyle factors related to cardiometabolic and reproductive health including socio-economic status, dietary intake, and physical activity [22,37,38].

In both studies, women who had a history of hysterectomy and/or ovariectomy or who were pregnant or lactating at the time of recruitment were excluded. Hormonal contraceptive use was not well captured in our cohorts for use as an exclusion criterion. The baseline characteristics between individuals with measured and unmeasured AMH levels are compared in Supplementary Table S2.

### Anthropometric and biochemical measurements

The collection and measurement of anthropometric, cardiometabolic and lifestyle-related data have been described in detail before [22,24,39]. Whole blood samples were collected for genotyping and serum biomarker measurements after an overnight fast [22,24,39].

Serum AMH levels were measured using manual enzyme linked immunosorbent assays (ELISA) from Ansh Labs (Webster, TX). In the 2002–03 family study, AMH levels were determined for 198 participants using the picoAMH ELISA assay; AMH levels for 14 women were determined with the Ultrasensitive (us)AMH/MIS ELISA assay. In the 2010 Soifua Manuia study, the picoAMH assay was used for women age ≥ 40 years old (*n* = 516) and the usAMH/MIS assay was used for women < 40 years old (*n* = 457) [21].The inter- and intraassay coefficients of variation were < 15%. The detection limit was 6 pg/mL and 0.08 ng/mL for the picoAMH assay and the usAMH/MIS assay, respectively. To harmonize measurements from the two different assays, the values from the picoAMH assay were rescaled to align them with values from the usAMH/MIS assay using this equation (Ansh Lab, insert AL124-i released on 2019-09-27, regression *R* ² = 0.99, *p* < 0.0001):

usAMH/MIS assay (ng/mL) = (picoAMH assay (pg/mL) + 50.66) / 0.92 / 1000

### Genotyping and imputation

Genotyping in 2002–03 family study was performed using the Global Screening Array-24 v.3.0 BeadChip (Illumina, CA, USA) with 644,880 SNVs including custom content pertinent to Samoans. For the 2010 Soifua Manuia study, 659,492 SNVs were genotyped genome-wide using Affymetrix 6.0 array. Quality control procedures were implemented for genotypes from both arrays following the guidelines outlined by Laurie et al. (2010). Detailed description of genotyping and quality control have been previously published [24,39].

Using a reference panel derived from 1,285 Samoan individuals with whole-genome sequencing [40], we performed imputation using minimac4 in both samples and removed variants with *R* ² < 0.3, yielding an additional 16,744,117 (2002–03 family study) and 15,633,124 (2010 Soifua Manuia study) SNVs [41].

### Ethical approval

Research protocols, informed consents and secondary analyses of both studies were approved by the Health Research Committee of the Samoan Ministry of Health and the Institutional Review Boards of Brown University as well as University of Cincinnati and University of Pittsburgh for the 2010 Soifua Manuia study and additionally the American Samoa Department of Health IRB for the 2002–03 family study [22,24,39]. All participants were informed about their rights verbally in Samoan by trained research staff before obtaining their written consent [22,24,39].

### Genome-wide association study

To account for AMH levels below the detection limit (2002–03 family study: *n* = 1; 2010 Soifua Manuia study: *n* = 169), we tested for association between genotype dosages and AMH levels using a Cox mixed-effects model as implemented in the R package {coxmeg} [42,43]. Since Cox regression is designed for right-truncated data, we used the reciprocal of the measured AMH levels [44]. We adjusted for fixed effects of centered age and centered age² (as well as polity for the 2002–03 family study) and for random effects of genetic relatedness using empirical kinship coefficients as estimated by PC-Relate [45]. Due to the genetic homogeneity of the sample [24], we did not adjust for principal components of ancestry.

The association results from both Samoan samples were meta-analyzed using a *p* value– based fixed-effect approach via METAL [46]. Before combining the two association studies, results from the 2002–03 family study and the 2010 Soifua Manuia study were filtered for minor allele frequency (MAF) ≥ 0.05 and MAF ≥ 0.01, respectively, to keep the minor allele count between the two samples close in range. SNVs with a *p* value ≤ 1 × 10⁻⁵ in the two pre–metaanalysis GWASs were tested for goodness of fit to Hardy–Weinberg equilibrium, and SNVs with *p* < 0.0001 were excluded from subsequent analyses. Conditional analyses to detect secondary signals were conducted by including the lead SNVs in each suggestively associated region as covariates in the original model.

Manhattan and QQ plots were created using the R package {fastman} [47]. Tests with *p* values ≤ 5 × 10⁻⁸ were considered genome-wide significant, and ≤ 1 × 10⁻⁵ were considered suggestive. Genomic positions are in Genome Reference Consortium Human Build 38 (hg38).

To refine the regions of associations and determine the most probable causal variant, each locus with a *p* value ≤ 1 × 10⁻⁵ was visualized with LocusZoom [48] and assigned a posterior probability via Bayesian fine-mapping with PAINTOR v2.1. [49] using functional annotation from the Ensembl Variant Effect Predictor (VEP, [50]) and RegulomeDB [51]. This fine-mapping was carried out in a region ±500 kb around each lead SNV, accounting for Samoan-specific linkage disequilibrium (LD) structure.

To identify gene sets and pathways that are enriched for variants relevant to AMH variation, GWAS meta-analysis summary statistics were processed by the Functional Mapping and Annotation (FUMA) v1.5.2 genetic associations pipeline [52] after converting the genomic positions from UCSC hg38 to genomic build UCSC hg19 via liftOver [53]. In addition to functional annotation, we used FUMA to carry out gene-based analysis via Multi-marker Analysis of GenoMic Annotation (MAGMA) [54] and preliminary expression quantitative trait loci (eQTL) analysis. Independent loci were sets of SNVs with a GWAS *p* value ≤ 1 × 10⁻⁵ and not in high LD (*r* ² < 0.6) with other SNVs with *p* value ≤ 1 × 10⁻⁵ on the same chromosome. Candidate SNVs for examination with FUMA were SNVs within ±500 kb of the lead SNV that had a GWAS *p* value < 0.05 and were in LD (*r* ² ≥ 0.6) with the lead SNVs. LD structure for FUMA analysis was calculated using all populations of the 1000G phase 3. Variants located outside the gene boundaries but relevant to the nearby gene are assigned to it by MAGMA. The annotation window size for proxy SNVs was set at 40 kb upstream and 10 kb downstream around each gene. The significance threshold for the gene-based test was set at *p* = 2.50 × 10⁻⁶ following Bonferroni correction for 20,000 tests. eQTL analysis interrogated whole blood, endocrine tissues (adrenal gland, hypothalamus, ovary, pancreas, pituitary, thyroid) and metabolic tissues (subcutaneous adipose tissue, liver) using data from GTEx v8 [55]. *p* values in the eQTL analyses were Bonferroni corrected based on the number of genes assessed.

### Known AMH loci

We examined our GWAS summary statistics for evidence of association of AMH levels with the reported lead SNVs in the eight known AMH loci (*TEX41* [2-144887307-A-G] [32,34]; *CDCA7* [2-173394597-C-T] [32]; *CTB-99A3.1* [5-146560687-G-A] [32]; *EIF4EBP1* [8-38015258-C-T] [34]; *BMP4* [14-53956049-G-T] [34]; *AMH* [19-2251818-T-C] [34]; *MCM8* [20-5967581-G-A] [31,32,34]; and *CHECK2* [22-28707610-T-C] [34]). A locus was considered replicated if the lead SNV was present in the meta-analysis, had a *p* value < 0.05, and had the same effect direction.

When the lead SNVs from these prior GWASs was absent from the meta-analysis, because the allele frequency was too low for inclusion in the meta-analysis, we also report the lead SNV within ±50 kb of the prior GWAS’s lead SNV if it had *p* < 0.05. For each of these nearby SNVs we also calculated a per-region Bonferroni-corrected significance threshold. The threshold was corrected for the number of independent SNVs in the region as calculated by simpleM [56].

### Transcriptome-wide association study

To assess the association of estimated gene expression levels based on genotypes with the phenotypes, we carried out a transcriptome-wide association study (TWAS) on meta-analysis summary statistics with the MetaXcan [57–59] suite of tools. We used the MASHR algorithm to predict gene expression, as it has demonstrated better performance than the Elastic Net algorithm [60]. We performed TWAS in whole blood, endocrine tissues (adrenal gland, hypothalamus, ovary, pancreas, pituitary, thyroid) and metabolic tissues (liver, subcutaneous adipose tissue) using SPrediXcan and combined the results for each gene from all tested single-tissue models into a single aggregate statistic using S-MultiXcan [58]. The *p* value for statistical significance (*p =* 2.61 × 10⁻⁶) is adjusted for the number of genes tested (*n* = 19,158).

### Colocalization analysis

Colocalization analyses were performed using fastENLOC [61–64]. For this, we converted the *z* scores from the meta-analysis to posterior inclusion probabilities (PIP) for causality via TORUS and colocalized 5,589,187 variants with eQTL data from GTEx within the nine issues listed above [65]. Precomputed GTEx multi-tissue annotations are available at https://github.com/xqwen/fastenloc [61–64].

## Results

The socio-demographic characteristics and AMH levels for the two samples are presented in Table 1, and AMH levels by age are depicted in Supplementary Figure S2.

**Table 1.**
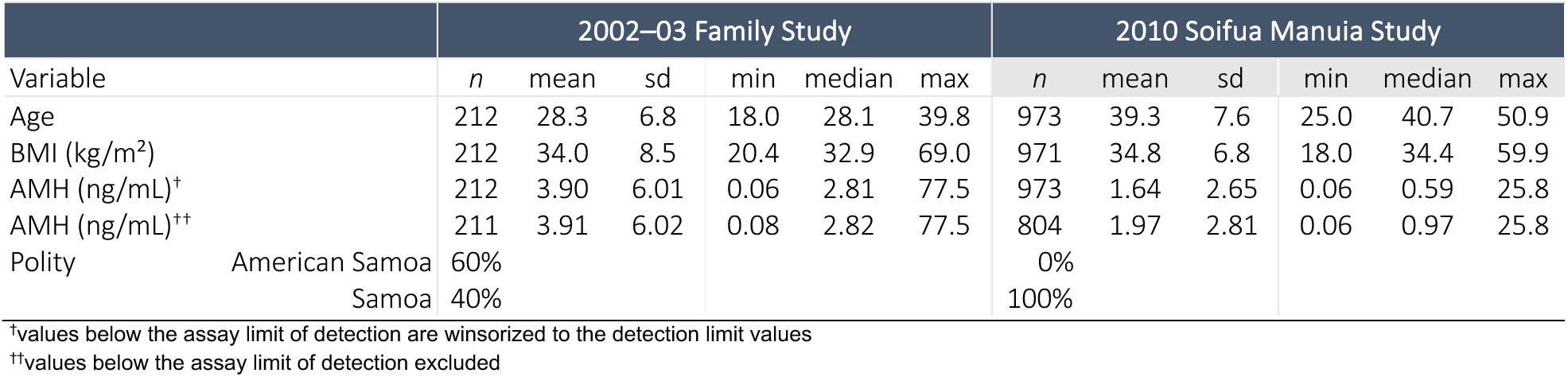
Sample demographics and characteristics for quantitative trait.

The GWAS of AMH levels had no genome-wide statistically significant associations (*p* ≤ 5 × 10⁻⁸) but eleven loci with *p* ≤ 1 × 10⁻⁵ were observed (Figure 1, Supplementary Figure S3, Table 2). The lead variants of each locus also had the highest posterior probability after fine-mapping for causal variants, except for the lead variant in *ARID3A* (Table 2). The quality score of all imputed lead variants were > 0.88. We confirmed via conditional analysis that no secondary signals were present in suggestive loci. We confirmed via conditional analysis that no secondary signals were present in suggestive loci. For each of the suggestive loci, the regional plots are shown in Supplementary Figure S4.

**Figure 1.**
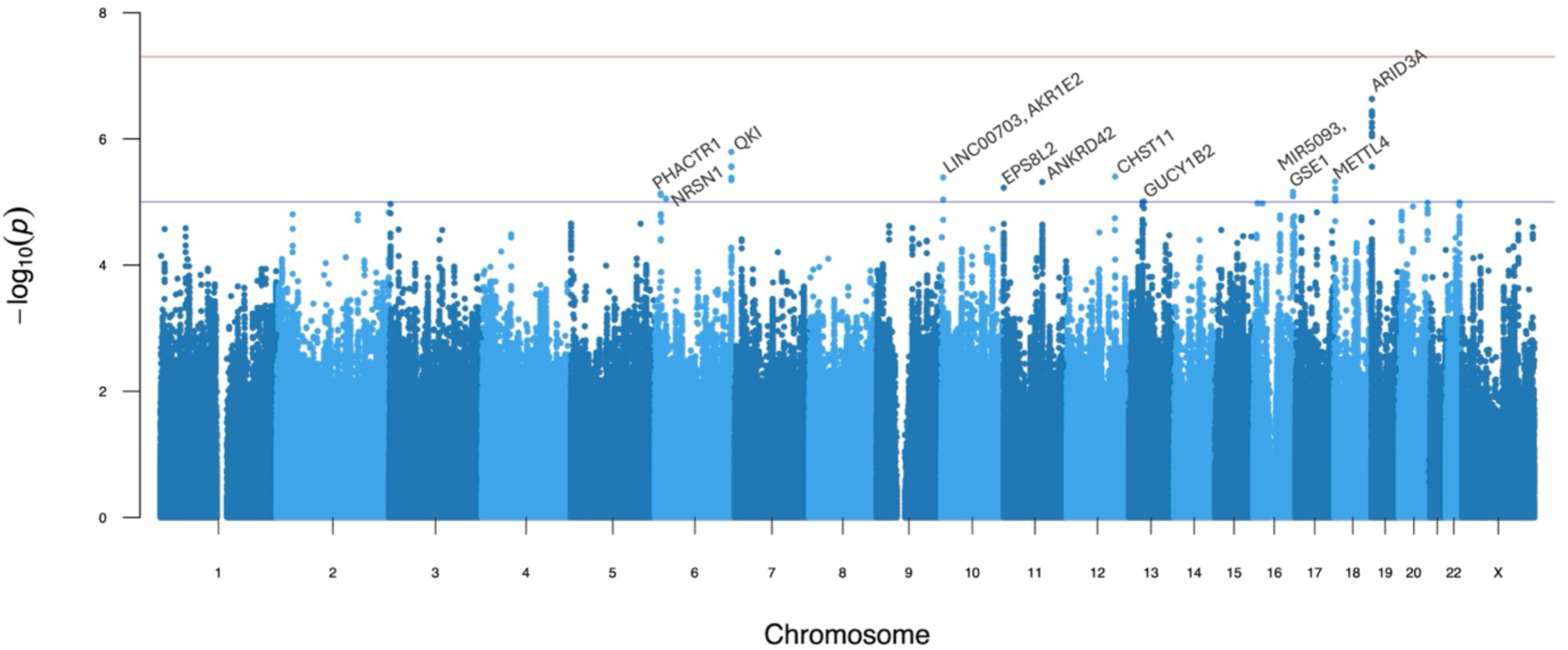
Manhattan plot for anti-Müllerian hormone levels. The solid red and blue lines denote the genome-wide significant and suggestive *p* value thresholds at *p* < 5 × 10⁻⁸ and *p* < 1 × 10⁻⁵, respectively. The peak SNV in each independent locus that surpassed the suggestive threshold is labeled with the nearby genes.

**Table 2.**
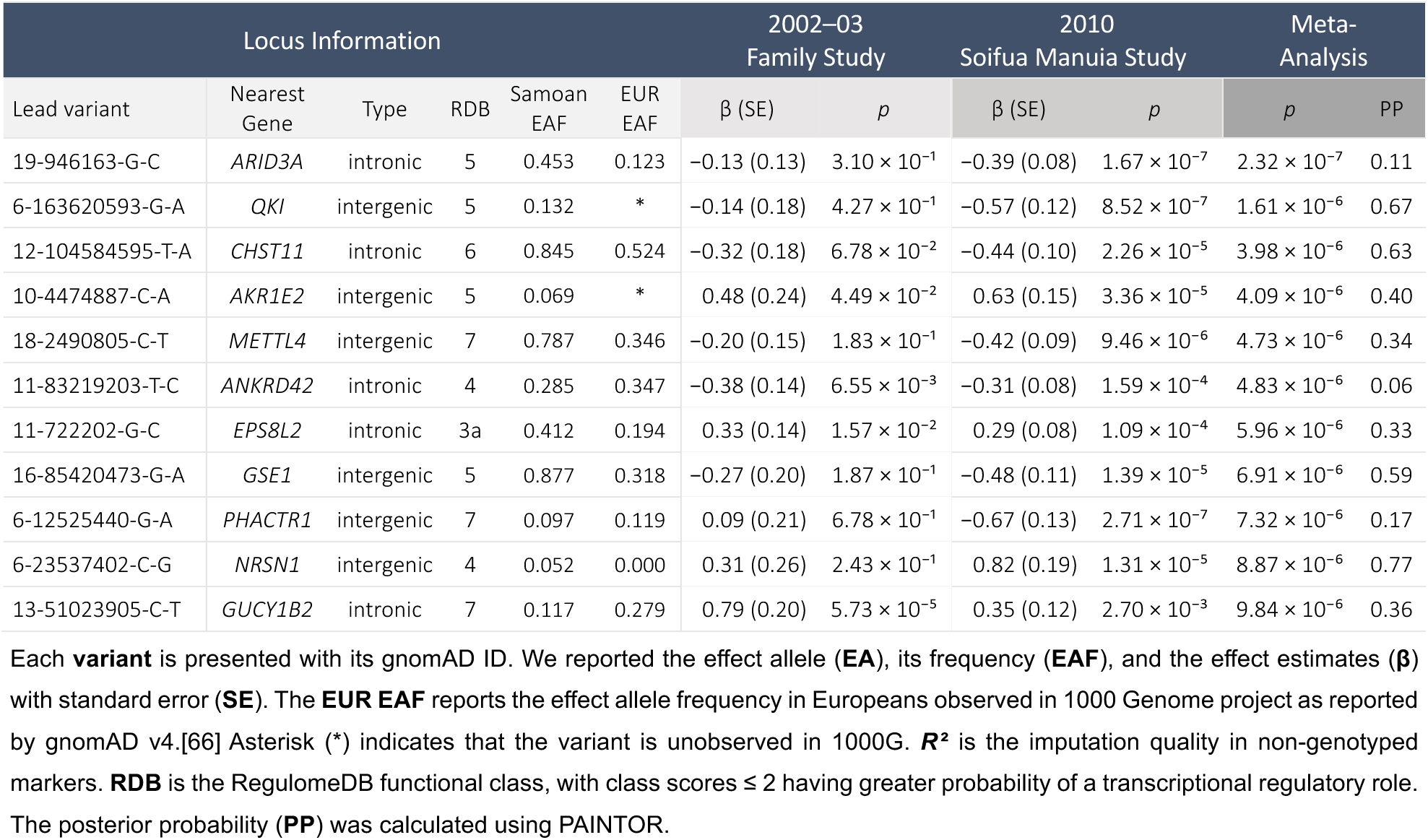
Meta-analysis results for AMH levels presenting independent SNVs with *p* value ≤ 1 × 10⁻⁵.

We replicated one of the eight known AMH loci: *EIF4EBP1* (Supplementary Table S3, Supplementary Figure S7 and Supplementary Figure S8). Of the seven known AMH loci with lead variants not replicated here, the lead variants in three (*MCM8*, *CHECK2* and *CTB99A3.1*) were ultra-rare in Samoans (MAF < 0.0001). Within ±50 kb of two unreplicated loci (*AMH* and *TEX41*), we detected two SNVs with a significant p-value (2-144839456-A-T [*p* = 0.0018] and 19-2250470G-A [*p* = 0.0006], respectively), however, they were not in LD with the known AMH loci.

The strongest GWAS association is in intron 3 of *ARID3A* at 19p13.3. The lead SNV, 19946163-G-C (*p* = 2.32 × 10⁻⁷), and nearby SNVs are presented in Figure 2. The alternate allele of the lead SNV was associated with lower AMH levels (Supplementary Figure S5 and Supplementary Figure S6). This locus also harbors the known age-at-menopause variant 19-950694-GA, observed in women of European ancestry, which is in high LD (*r* ² = 0.79) with the lead AMH variant in this locus in this study [67,68]. Fine-mapping identified 19-982128-A-G, upstream of *WDR18*, as the most probable causal variant with a posterior probability (PP) of 0.34. The lead AMH GWAS variant, 19-946163-G-C, (PP = 0.11) and age-at-menopause variant 19-950694-GA (PP = 0.02) were the top two eQTLs affecting *ARID3A* expression in thyroid tissue (*p* = 5.75 × 10⁻⁷ and *p* = 3.69 × 10⁻⁷, respectively, both with an FDR = 2.60 × 10⁻¹⁰).

**Figure 2.**
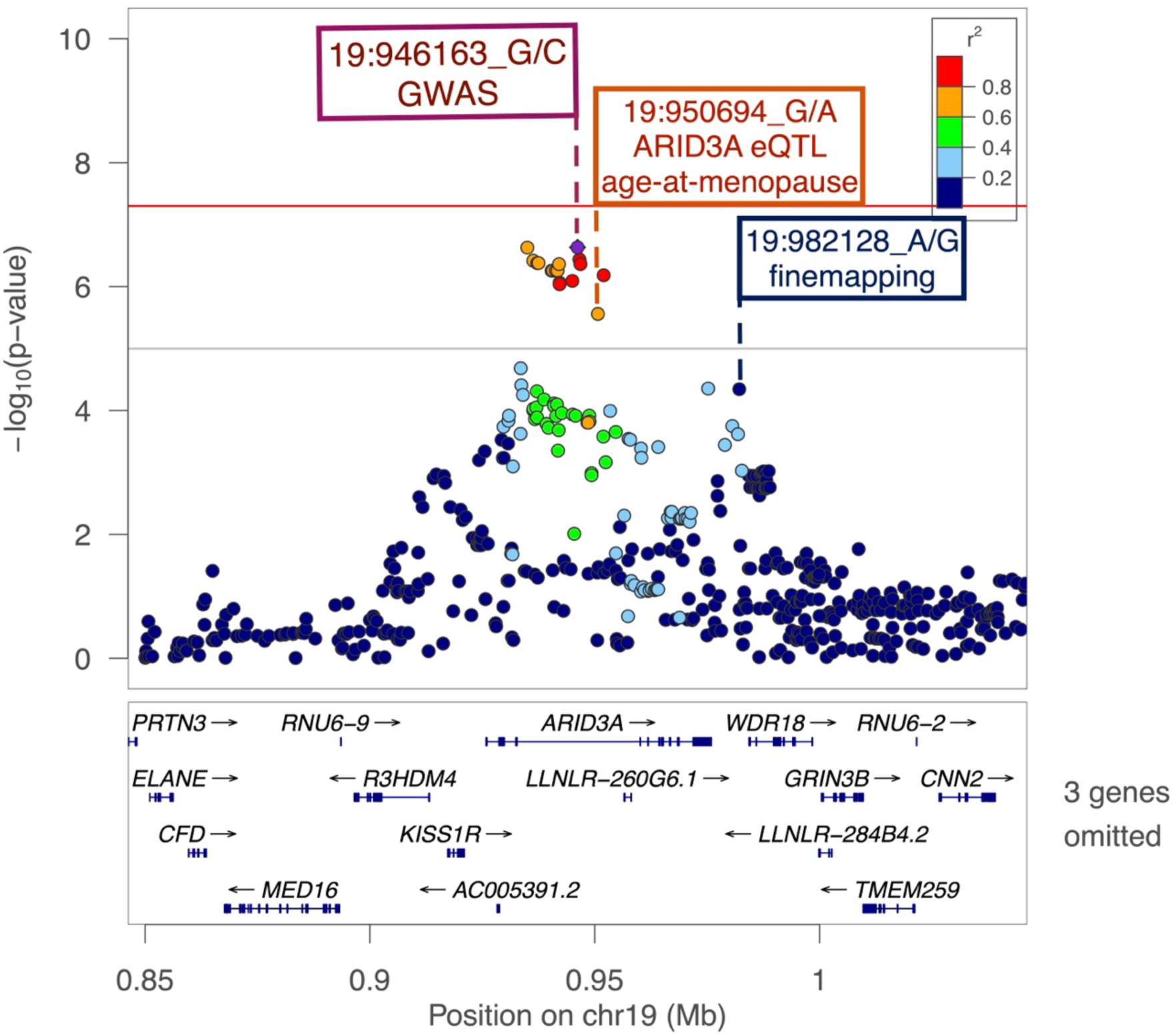
Regional plot for *ARID3A* locus. 19-946163-G-C, the lead SNV, is a purple diamond. The color of all other SNVs reflects LD with the lead SNV as calculated in the Samoan samples. The solid red and grey line indicate the significance and suggestive thresholds at *p* < 5 × 10⁻⁸ and *p* < 1 × 10⁻⁵, respectively.

Gene-based analysis identified five significant associations with AMH levels: *ARID3A* (*p =* 5 × 10⁻¹⁰) and nearby *R3HDM4* (*p =* 1.47 × 10⁻⁹) as well as *P2RX6* (*p =* 1.15 × 10⁻⁶), *AC002472.1* (*p =* 8.30 × 10⁻⁷), and *PTPRB* (*p =* 2.47× 10⁻⁶) (Supplementary Figure S9).

There were seven transcriptome-wide significant genes observed in the TWAS of whole blood, endocrine tissues (adrenal gland, hypothalamus, ovary, pancreas, pituitary, thyroid), and metabolic tissues (liver, subcutaneous adipose tissue) in association with AMH levels. The strongest association of the seven was *GINS2*. *METTL4* was not only transcriptome-wide significance but also suggestively associated with AMH levels. The TWAS results are presented in Figure 3 and Supplementary Table S4.

**Figure 3.**
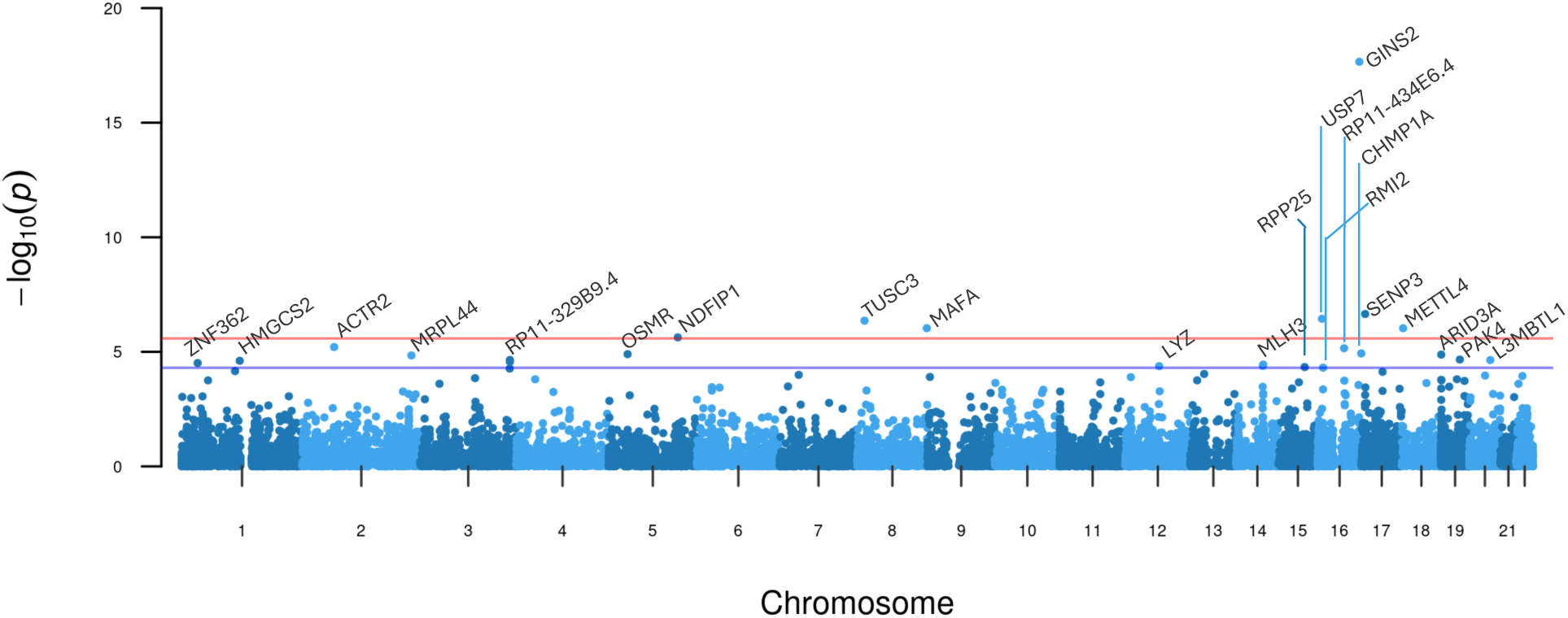
Manhattan plot of TWAS results for AMH. The solid red and blue lines denote the transcriptome-wide significant and suggestive *p* value thresholds at *p* < 2.6 × 10⁻⁶ and *p* < 1 × 10⁻⁵, respectively. The genes that surpassed the suggestive threshold are annotated.

We conducted a colocalization analysis using summary statistics from the GWAS and TWAS. In SNV-level and gene-level colocalization analysis, *ARID3A* had a locus-level colocalization probability (LCP) of 0.34 in the liver. All other colocalization probabilities for other genes and tissues were below the threshold of 0.30.

## Discussion

Here we report the first study examining association between genetic variants and AMH levels in women from a Polynesian population. We identified eleven novel suggestive loci via GWAS (*ARID3A, QKI, CHST11, AKR1E2, METTL4, ANKRD42, EPS8L2, GSE1, EDN1, NRSN1,* and *GUCY1B2*) and seven significant genes via TWAS (*GINS2, SENP3, USP7, TUSC3, MAFA, METTL4*, and *NDFIP1*). Additionally, we replicated the known AMH locus *EIF4EBP1* previously detected in women of European ancestry [34]. Among these findings, three genomic risk loci— *ARID3A, GSE1* and nearby *GINS2,* and *METTL4*—had associations in both GWAS and TWAS and are biologically plausible.

Our key GWAS finding was association of AMH with variants in the gene encoding AT-rich interaction domain 3A (*ARID3A*), a member of a family of proteins regulating chromatin binding. It was a also significant locus in gene-based analysis and was suggestively significant in the TWAS. The lead variant in *ARID3A* is located 4.5 kb upstream from a known age-at-menopause variant [67,68] and the two are in high LD. The biological link between AMH levels (a marker of ovarian follicle reserve) and age-at menopause is well recognized, as AMH testing is used to predict time-to-menopause in late reproductive age women in clinical practice [69,70]. Notably, both the lead *ARID3A* GWAS variant and the known age-at-menopause variant are positioned within GeneHancer [71] regulatory element GH19J000942, a target of which is the gene encoding KISS1 (kisspeptin) receptor (*KISS1R*). While there is not strong statistical evidence of gene-based or TWAS associations for *KISS1R* itself in this study, that does not rule it out as having a causal role in this locus. Kisspeptin/KISS1R signaling is pivotal in folliculogenesis [72]. Increased ovarian kisspeptin levels hinder the transition of primary follicle into antral stage by inhibiting the *FSHR* expression and increasing the AMH levels [72]. Observations in mice with conditional ablation of *Kiss1r* in oocytes suggest that the resulting deregulation may lead to premature ovarian failure [73]. Although the lead variant is in an intron of *ARID3A*, should this variant be causal or in LD with the causal variant, it may be acting through an effect on *KISS1R*.

In the TWAS, *GINS2* was the most significant gene and is within the *GSE1* locus on 16q24.1, which was suggestively significant in the GWAS. *GINS2* is highly conserved among eukaryotes and encodes one of the essential subunits that form the tetrameric Go-Ichi-Nii-San (GINS) complex, which has a key role in DNA replication [74,75]. *GINS2* is downregulated in ovaries of old-aged rhesus monkeys compared to young- and middle-aged ones and in atretic bovine ovarian follicles compared to the healthy ones [76,77]. *GSE1*, an epigenetic regulator and a known oncogene, may also have a biological connection to AMH levels, as it has highest expression in the pituitary and the ovary [55,78]. The expression of *Gse1* in the primordial follicles is downregulated in estrogen receptor β (ERβ) knockout mice [79]. Loss of ERβ activates follicle growth and leads to early depletion of the ovary reserve, suggesting a regulatory role for *Gse1* in folliculogenesis [79]. AMH stimulates gonadotropin-releasing hormone (GnRH) expression in its role in the hypothalamic-pituitary-gonadal hormonal axis, and therefore, it is notable that *Gse1* is highly enriched in GnRH neurons upon gonadectomy [80,81].

The *METTL4* locus on 18p11.32 includes *METTL4* and *NDC80* and was identified by both GWAS and TWAS analyses (Supplementary Figure S4J). *METTL4* encodes methyltransferase 4, which is responsible for the adenine methylation involved in regulating RNA-splicing [82]. Epigenetic regulation via N^6^-methyladenosine modifications plays an active role in response to environmental stressors (hypoxia, starvation, toxicants, etc.) and has been reported to be a relevant mechanism in the development of PCOS and premature ovarian insufficiency [83–87]. While *METTL3*, a paralog of *METTL4*, has been implicated in follicle development and fertility by regulating the stability of oocyte meiotic maturation-related transcripts in mice, not much is known about the role of *METTL4* in female reproduction [82,88,89]. Nearby *NDC80* encodes a component of the nuclear division cycle 80 kinetochore complex [90,91]. In mouse oocytes, this complex partakes in initiating oocyte maturation by enabling the spindle assembly required for G2/M transition by stabilizing cyclin B2 levels [91–95]. Consequently, the dormant primordial follicles arrested since birth at G2/M resume meiosis [91–94]. Association of variants near *METTL4* may indicate that genes that control the G2/M transition have a biological connection to modulation AMH levels.

We replicated one of the eight known AMH loci—intergenic variant 8-38015258-C-T (rs10093345) near *EIF4EBP1*. This SNV was also significantly associated with age-at-menopause in the UK Biobank [96]. Notably, lead variants in four known AMH loci—*AMH*, *CHECK2*, *CTB-99A3.1*, and *MCM8—*are low-frequency or rare variants in Europeans; these same variants in Samoans are either rare with low Samoan-specific imputation quality (*R* ² < 0.10) or are ultrarare (MAF < 0.0007). Such population-specific susceptibility loci highlight differences in AMH genetic architecture between Samoan individuals and individuals of European ancestry and highlight the importance of diversifying study populations. Recently, Moolhuijsen et al. investigated whether *AMH* promoter variation affects serum AMH levels in PCOS patients of Northern European ancestry and observed an association between rs10406324 (19-2249113-G-A) and lower AMH levels that was independent of follicle count and other PCOS markers [33]. Similar findings were also observed in normo-ovulatory women [32]. This suggests that genetic factors can contribute to variation in AMH levels independent from follicle count and/or PCOS status. In this study, rs10406324 could not be analyzed due to low MAF (< 0.005) and poor imputation quality (*R* ² < 0.10).

## Strengths and limitations

While our sample was small compared to other published GWASs, we were uniquely positioned to identify variants that may be rare in other populations but common in Samoans due to population founder effects. Additionally, the genetic homogeneity of the Soifua Manuia [24] sample could result in better power to detect variants associated with AMH levels.

Measuring AMH levels in older women is a challenge, as AMH levels fall below detectable ranges during perimenopause. We addressed this challenge by employing two AMH assays from Ansh Lab: picoAMH and ultra-sensitive AMH/MIS ELISA kits for older women and younger women, respectively. These assays utilize the same antibodies and calibrators, with picoAMH extending coverage for the lower ranges of the standard curve [97]. To reduce the heterogeneity, we used the conversion factor from Ansh Lab to harmonize the AMH levels between the two age groups. Additionally, we robustly accounted for the AMH values below detectable limits in our GWASs leveraging the Cox’s proportional hazards regression model via coxmeg.

For GWAS findings, although we cannot entirely rule out the possibility of false positive (FP) results and acknowledge that a plausible story for those can be made easily [98], we aimed to mitigate this by prioritizing the findings with evidence from both GWAS and TWAS analyses. Furthermore, while we identified several significant TWAS associations, absence of Samoan-specific eQTLs and limited representation of diverse populations in GTEx could have led to reduced power and detection of fewer eQTL associations.

## Conclusion

Overall, eleven loci were suggestively associated with AMH levels in a meta-analysis of Samoan women, several of which have plausible links to ovarian function/folliculogenesis (*KISS1R, GINS2,* and *NDC80*). This study provides valuable insights into the genetic variation of AMH in Samoan women and replicates the previously detected association of *EIF4BP1* with AMH levels. The putative novel findings in this study will need to be validated in additional larger studies. The identification of variations affecting AMH levels, such as seen in our findings, may also improve understanding of the biological underpinnings of AMH-related reproductive traits such as ovarian function, age at menopause, premature ovarian failure, and PCOS. Eventually, these findings may contribute to the development of screening tools measuring the genetic susceptibility for AMH-related traits.

## Authors’ role

Z.E.-Y. conceptualized the study, performed the analyses, created visual representations of the results, contributed to the interpretation of the results and took the lead in writing the manuscript.

M.K. and J.Z.Z. imputed the genotypes. G.L.-M. conducted the biomarker analyses. G.L.-M., T.N., S.V., N.L.H., S.T.M., D.E.W. and N.L.H. provided resources and curated data. J.C.C. set up the analysis pipeline and supported methodological approach. D.E.W. and R.L.M. supervised the project, provided statistical and subject matter expertise, and helped develop the study design.

J.C.C., G.L.-M., T.N., S.V., N.L.H., S.T.M., D.E.W. and R.L.M. contributed to interpretation of the results and revision of the article. All authors read and approved the final manuscript.

## Data Availability Statement

The full AMH GWAS summary statistics will made available after publication through the GWAS catalog (https://www.ebi.ac.uk/gwas/).

## Acknowledgements

The authors thank all study participants for their participation and contribution to this research. We acknowledge the assistance of the Samoa Ministry of Health and the Samoa Bureau of Statistics for their guidance and support in the conduct of this study. We thank the local village officials for their help and the participants for their generosity. We further acknowledge helpful comments of the TOPMed Reproductive Health Working Group.

## Funding

This work was funded by NIH Grants R01-HL093093 (PI: S.T.M.), R01-HL133040 (PI: R.L.M.), and T90-DE030853 (PI: C.S. Sfeir). Molecular data for the Trans-Omics in Precision Medicine (TOPMed) Program was supported by the National Heart, Lung and Blood Institute (NHLBI). Genome sequencing for the Soifua Manuia study, labeled as “NHLBI TOPMed: Genome-wide Association Study of Adiposity in Samoans” (phs000972) in the dbGaP, was performed at the Northwest Genomics Center (HHSN268201100037C) and the New York Genome Center (HHSN268201500016C). Core support including centralized genomic read mapping and genotype calling, along with variant quality metrics and filtering were provided by the TOPMed Informatics Research Center (3R01-HL117626-02S1; contract HHSN268201800002I). Core support including phenotype harmonization, data management, sample-identity QC, and general program coordination were provided by the TOPMed Data Coordinating Center (R01-HL120393; U01-HL120393; contract HHSN268201800001I). The content is solely the responsibility of the authors and does not necessarily represent the official views of the National Institutes of Health.

## Conflict of interest

None declared.

## Notes

### Competing Interest Statement

The authors have declared no competing interest.

### Funding Statement

Research reported in this publication was funded by NIH grants R01HL093093 (PI: S.T.M.), R01HL133040 (PI: R.L.M.), and T90DE030853 (PI: C.S. Sfeir). Molecular data for the Trans-Omics in Precision Medicine (TOPMed) Program was supported by the National Heart, Lung and Blood Institute (NHLBI). The content is solely the responsibility of the authors and does not represent the official views of the National Institutes of Health.

### Author Declarations

Research protocols, informed consents and secondary analyses of both studies were approved by the Health Research Committee of the Samoan Ministry of Health and the Institutional Review Boards of Brown University as well as University of Cincinnati and University of Pittsburgh for the 2010 Soifua Manuia study and additionally the American Samoa Department of Health IRB for the 2002–03 family study.

