## Supplementary for "A genome-wide association study of anti-Müllerian hormone (AMH) levels in Samoan women"

## to

**Supplementary Tables**

### Abstract in Samoan

A practice of our research group is to make an abstract that has been translated to Samoan available. At this time, only English language text is allowed by medRxiv. A Samoan translation is available upon request from.

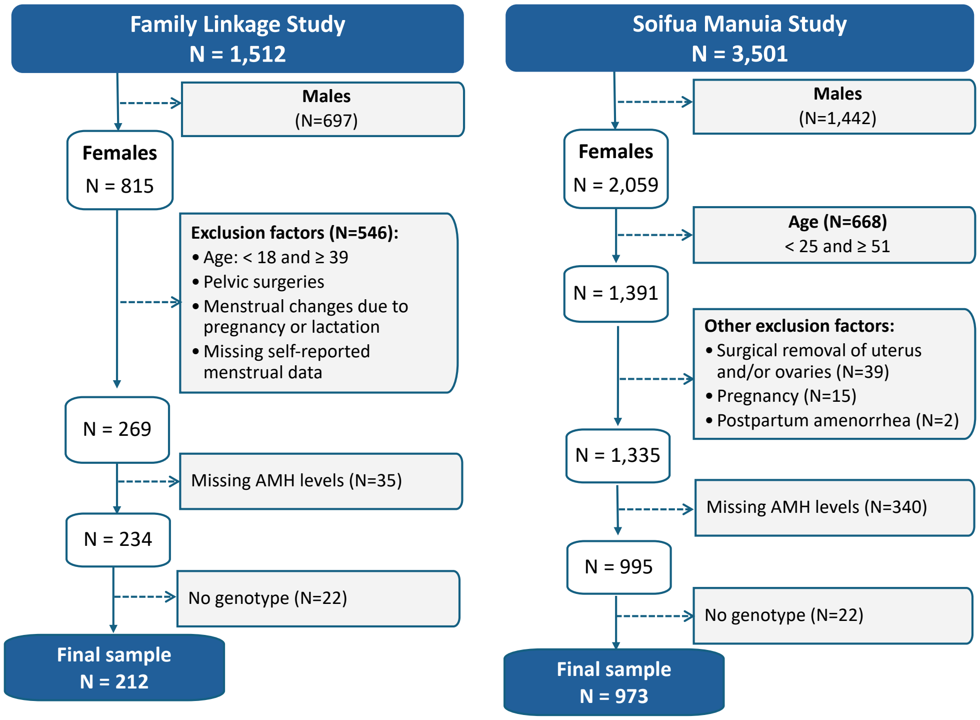

**Supplementary Figure S1 Flow chart diagrams showing the sample selection for Study Sample 1 (left) and for Study Sample 2 (right).**

**Supplementary Table S1** Statistical comparison of baseline characteristics between individuals with measured and unmeasured serum AMH levels.

| | Family Linkage Study | | | | | Soifua Manuia Study | | | | |
|---|---|---|---|---|---|---|---|---|---|---|
| | AMH | | | | | AMH | | | | |
| Characteristics | Unmeasured N = 35 | | Measured N = 234 | | p-value[1] | Unmeasured N = 340 | | Measured N = 995 | | p-value[1] |
| | N | mean (sd) | N | mean (sd) | | N | mean (sd) | N | mean (sd) | |
| Age | 35 | 30.26 (6.59) | 234 | 28.41 (6.76) | 0.11 | 340 | 36.23 (6.38) | 995 | 39.25 (7.64) | <0.001 |
| BMI | 35 | 34.62 (6.25) | 234 | 34.17 (8.55) | 0.5 | 338 | 34.10 (5.97) | 993 | 34.70 (6.83) | 0.4 |
| Glucose | 31 | 85.74 (13.24) | 234 | 90.79 (34.17) | > 0.9 | 191 | 91.32 (31.87) | 942 | 98.80 (46.04) | 0.081 |
| Insulin | 31 | 8.78 (9.19) | 234 | 15.48 (19.64) | 0.011 | 191 | 17.38 (14.18) | 941 | 17.06 (17.10) | 0.5 |
| HDL | 31 | 41.72 (9.28) | 234 | 44.90 (9.63) | 0.11 | 191 | 47.20 (11.67) | 942 | 47.15 (11.13) | 0.7 |
| LDL | 31 | 106.98 (27.17) | 233 | 111.28 (29.16) | 0.4 | 191 | 118.89 (28.30) | 941 | 124.16 (30.46) | 0.013 |
| NetTG | 31 | 124.74 (82.34) | 234 | 104.20 (60.68) | 0.4 | 191 | 97.14 (52.20) | 942 | 106.70 (91.59) | 0.049 |
| Cholesterol | 31 | 173.65 (30.21) | 234 | 176.92 (32.07) | 0.6 | 191 | 185.52 (32.19) | 942 | 192.33 (33.38) | 0.002 |

[1] Wilcoxon rank sum test

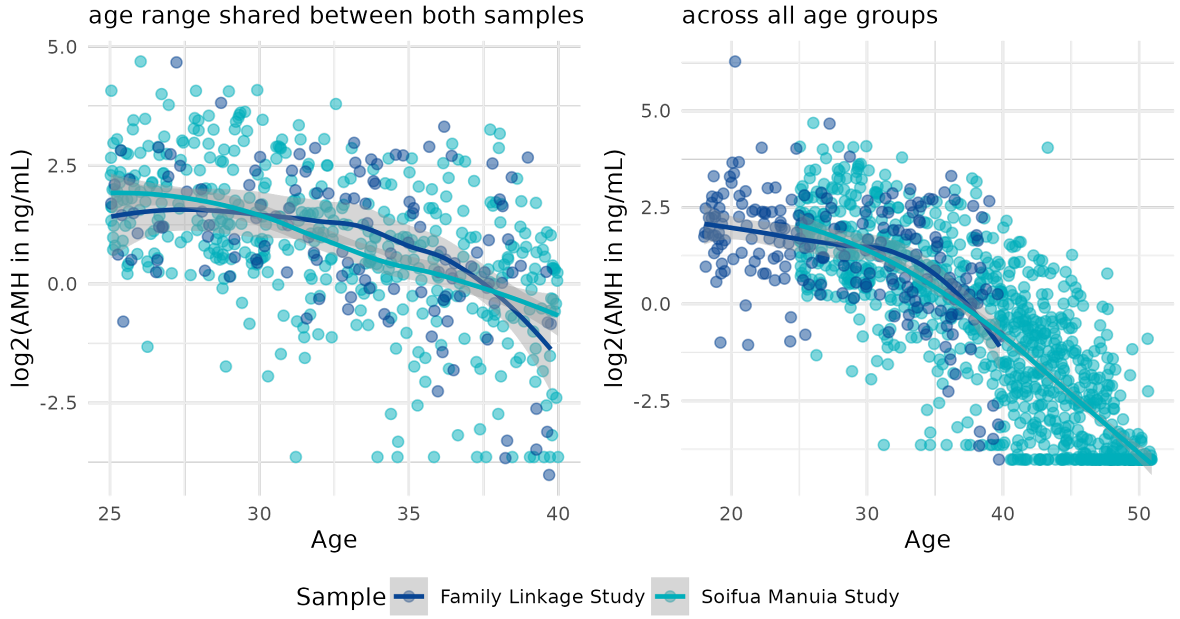

**Supplementary Figure S2 AMH levels by age in both samples.**

The $log_2$-transformed AMH levels by age are compared in overlapping (left) vs. across all (right) age ranges with smoothed loess curve.

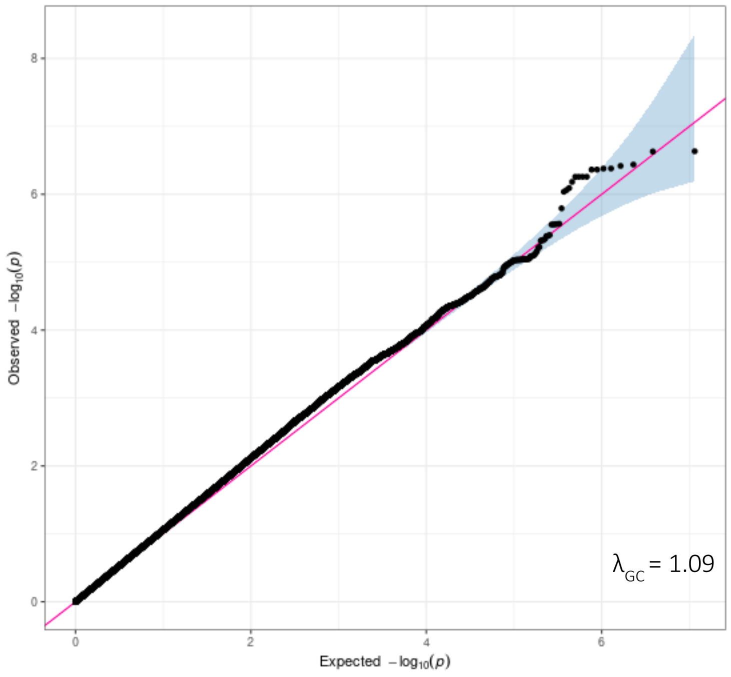

**Supplementary Figure S3 Quantile–quantile plot for AMH genome-wide association meta-analysis**

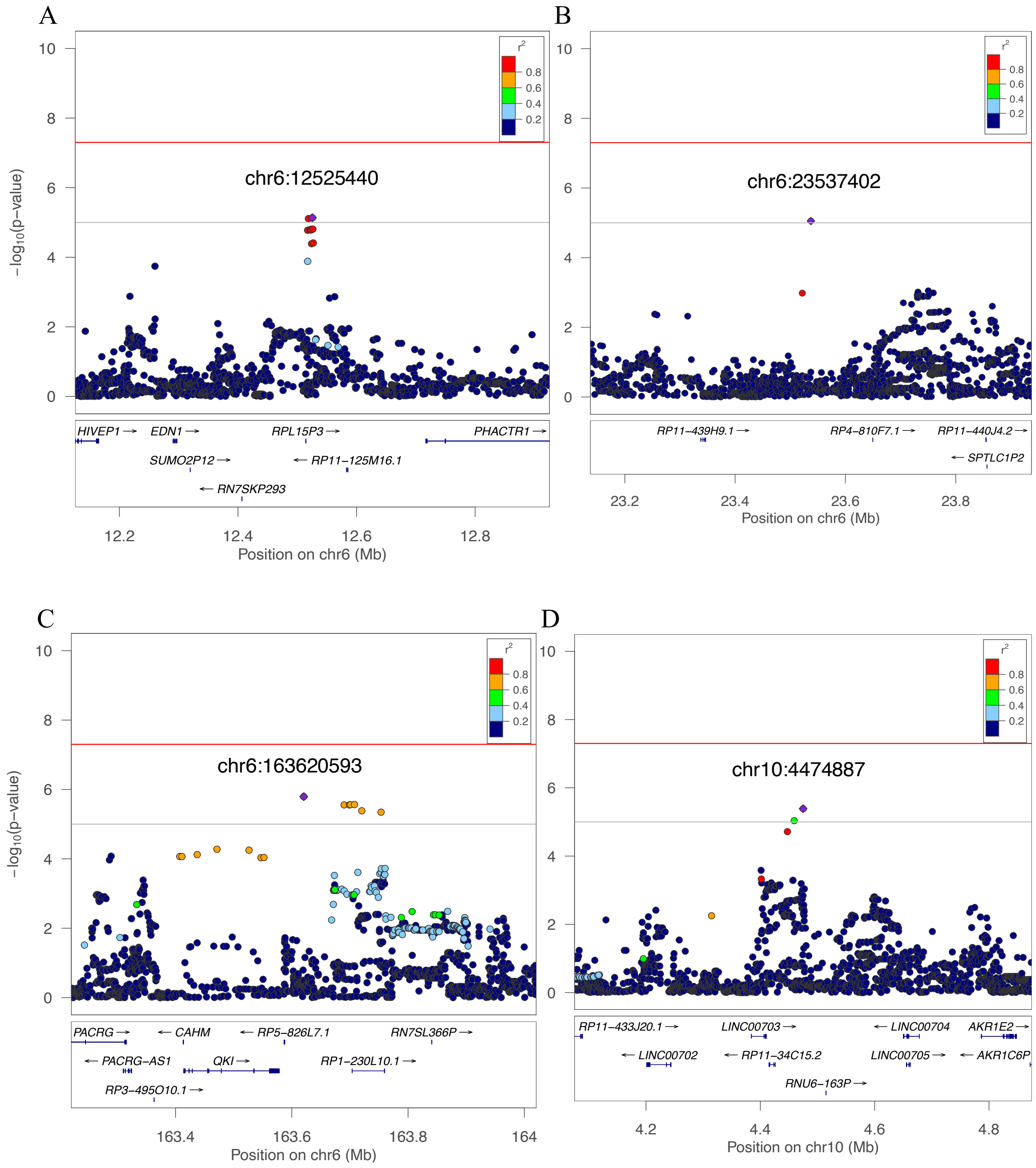

**Supplementary Figure S4 Regional plots for genome-wide association on AMH**

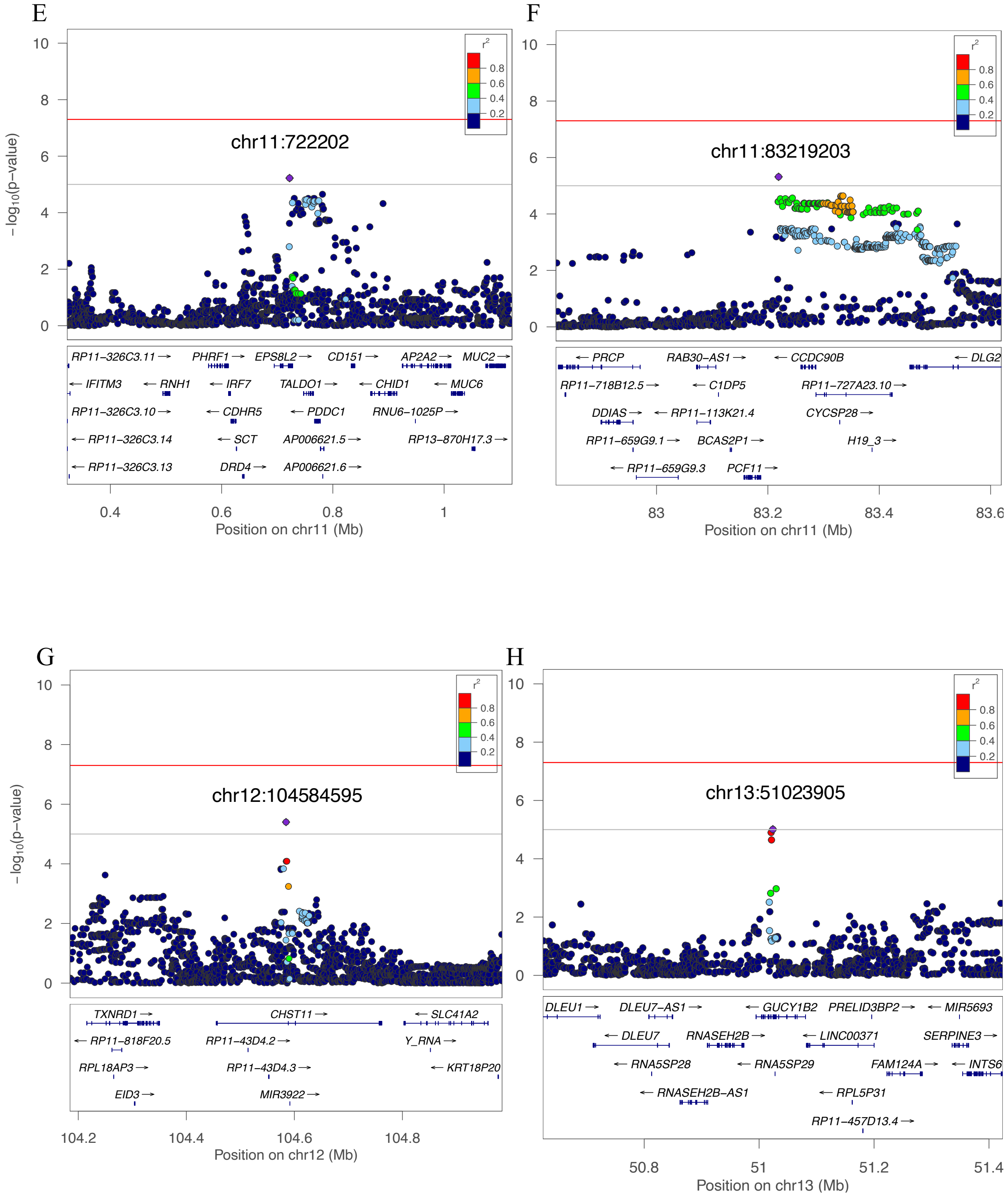

**Supplementary Figure S4 (cont'd). Regional plots for genome-wide association on AMH**

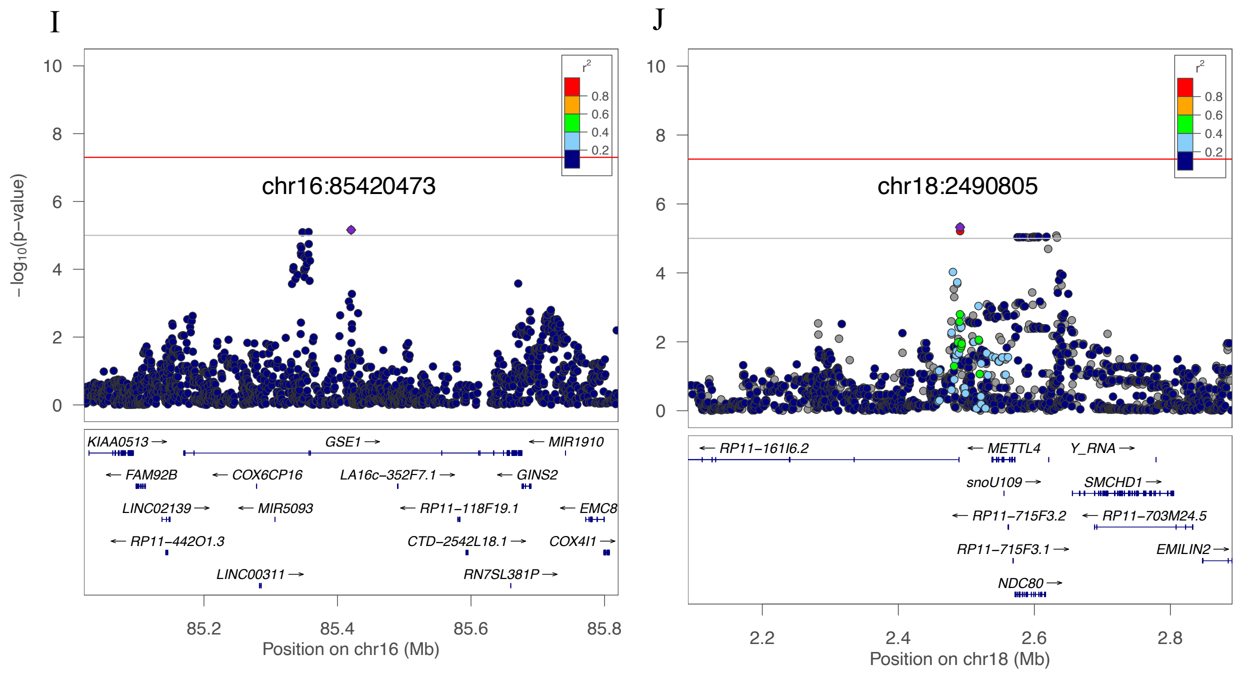

**Supplementary Figure S4 (cont'd). Regional plots for genome-wide association on AMH**

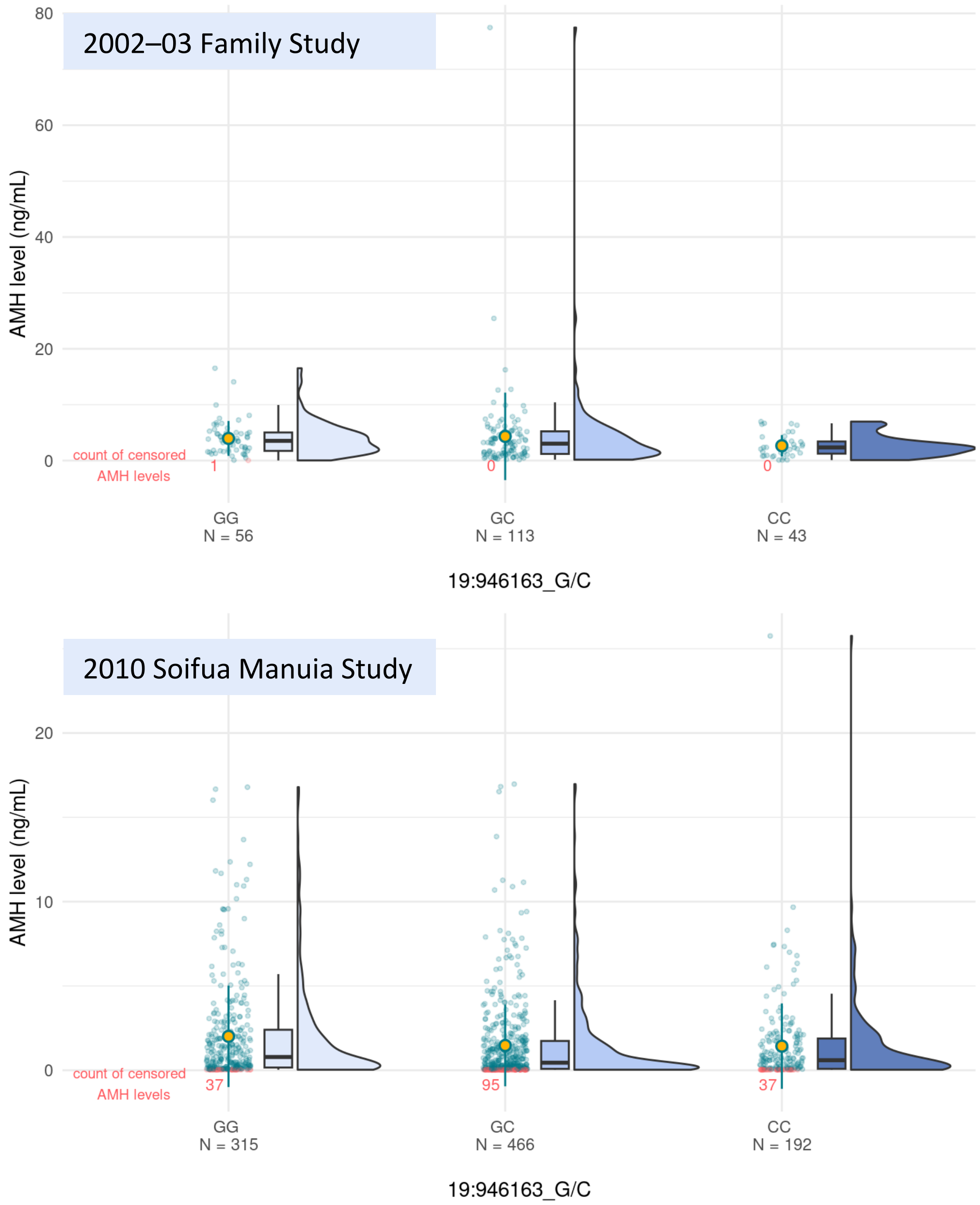

**Supplementary Figure S5 AMH levels for the lead variant 19-946163-G-C at *ARID3A* in 2002–03 Family Study (top) and 2010 Soifua Manuia Study (bottom)**

The mean (orange dot) and median (center line of box plot) AMH levels in ng/mL are shown by genotype. The AMH levels below detectable limits (red dots) are show with values set to the limit of detection divided by 2.

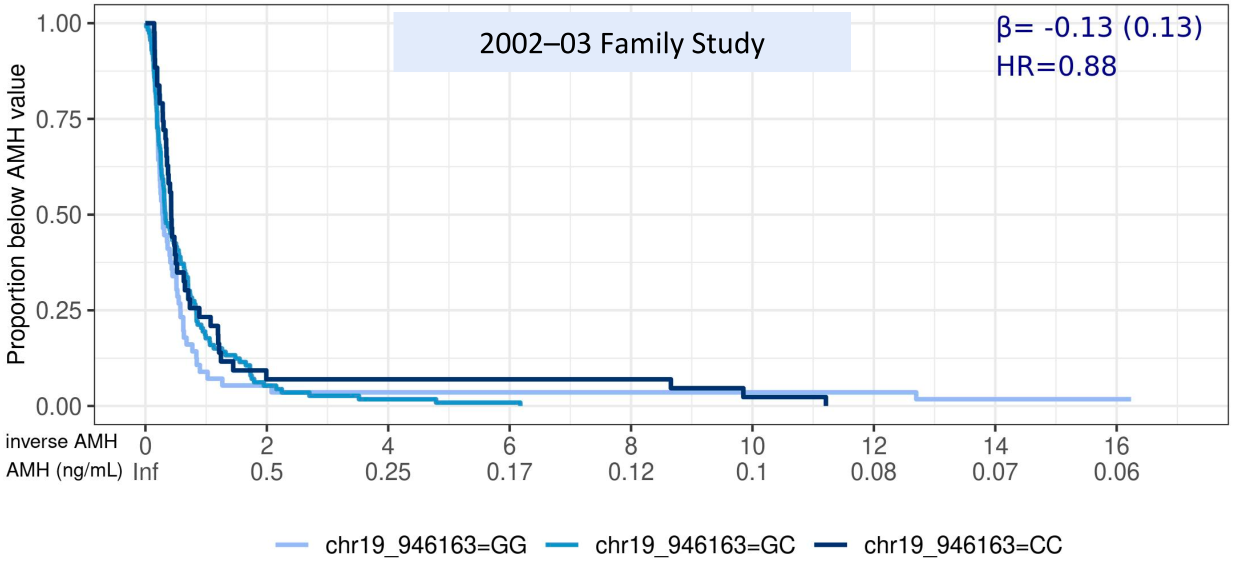

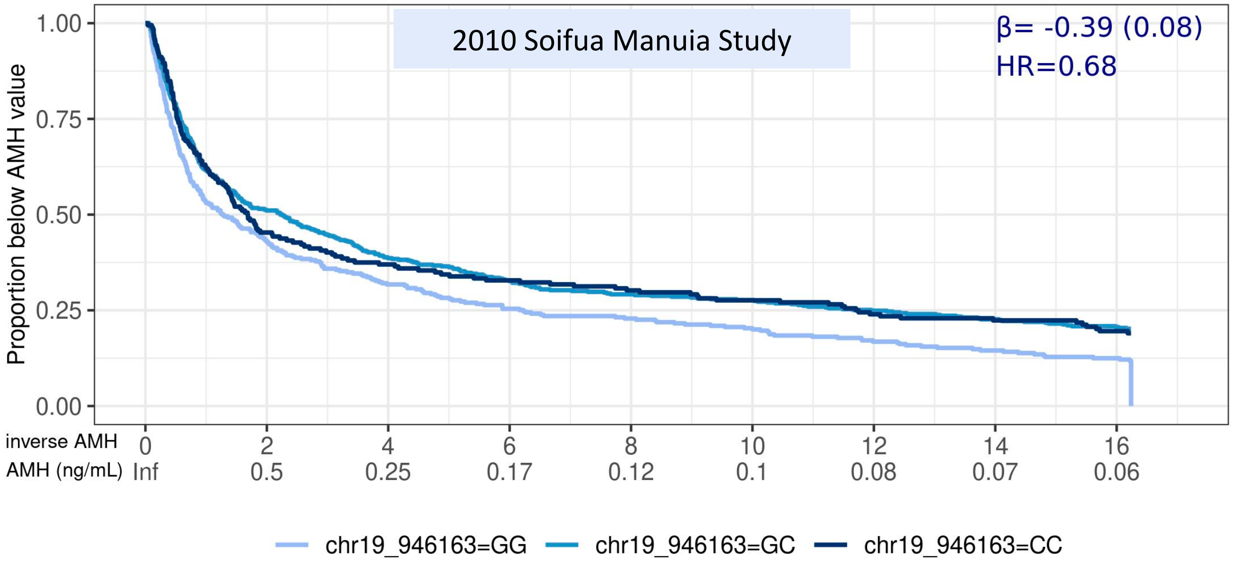

**Supplementary Figure S6 AMH levels by genotype strata for 2002–03 Family Study (top) and 2010 Soifua Manuia Study (bottom) using survival curve**

The inverse (or reciprocal) of the AMH level is shown in the *x* axis with the corresponding AMH levels in ng/mL beneath.

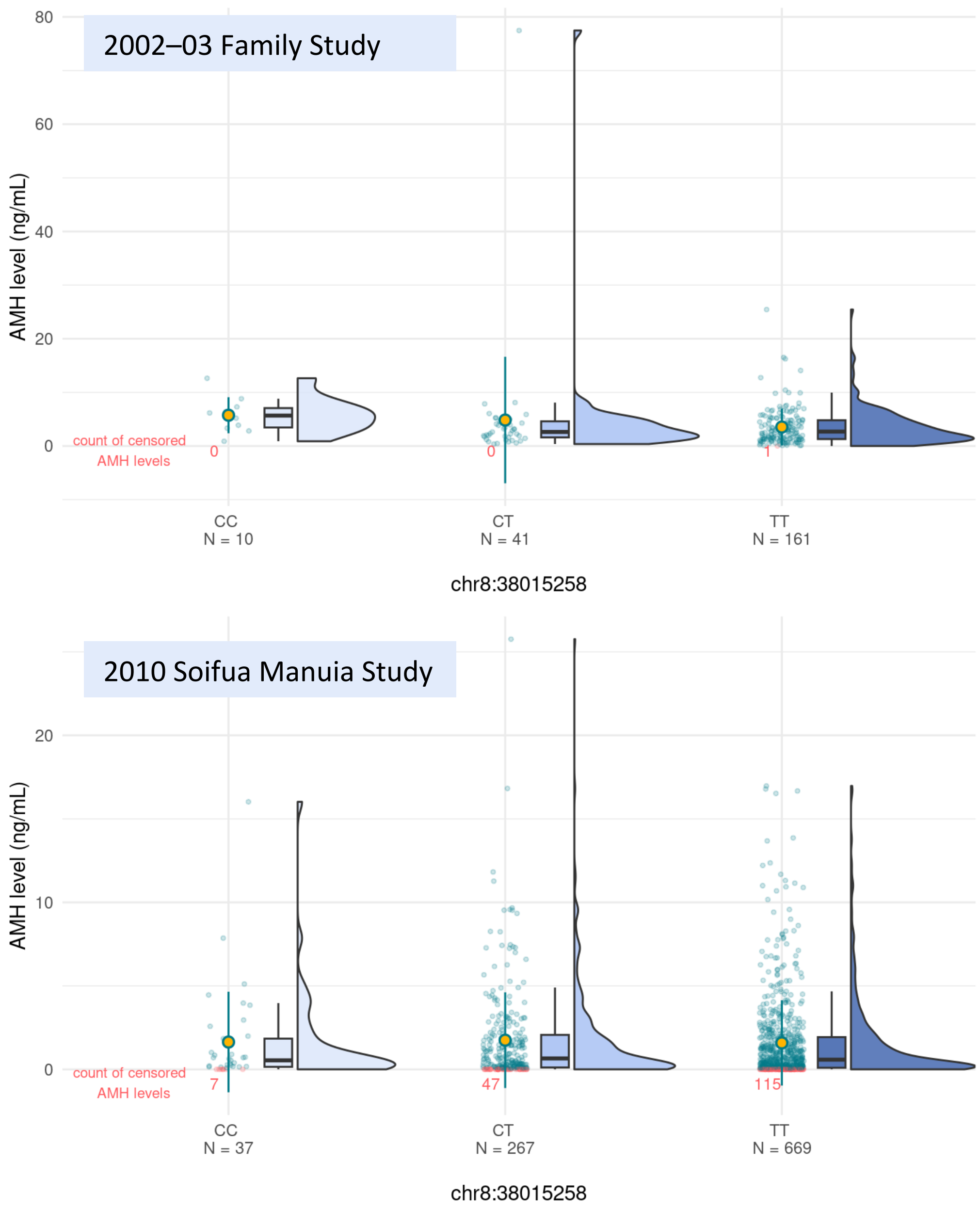

**Supplementary Figure S7 AMH levels for the lead variant 8-38015258-C-T at *EIF4EBP1* in 2002–03 Family Study (top) and 2010 Soifua Manuia Study (bottom)**

The mean (orange dot) and median (center line of box plot) AMH levels in ng/mL are shown by genotype. The AMH levels below detectable limits (red dots) are show with values set to the limit of detection divided by 2.

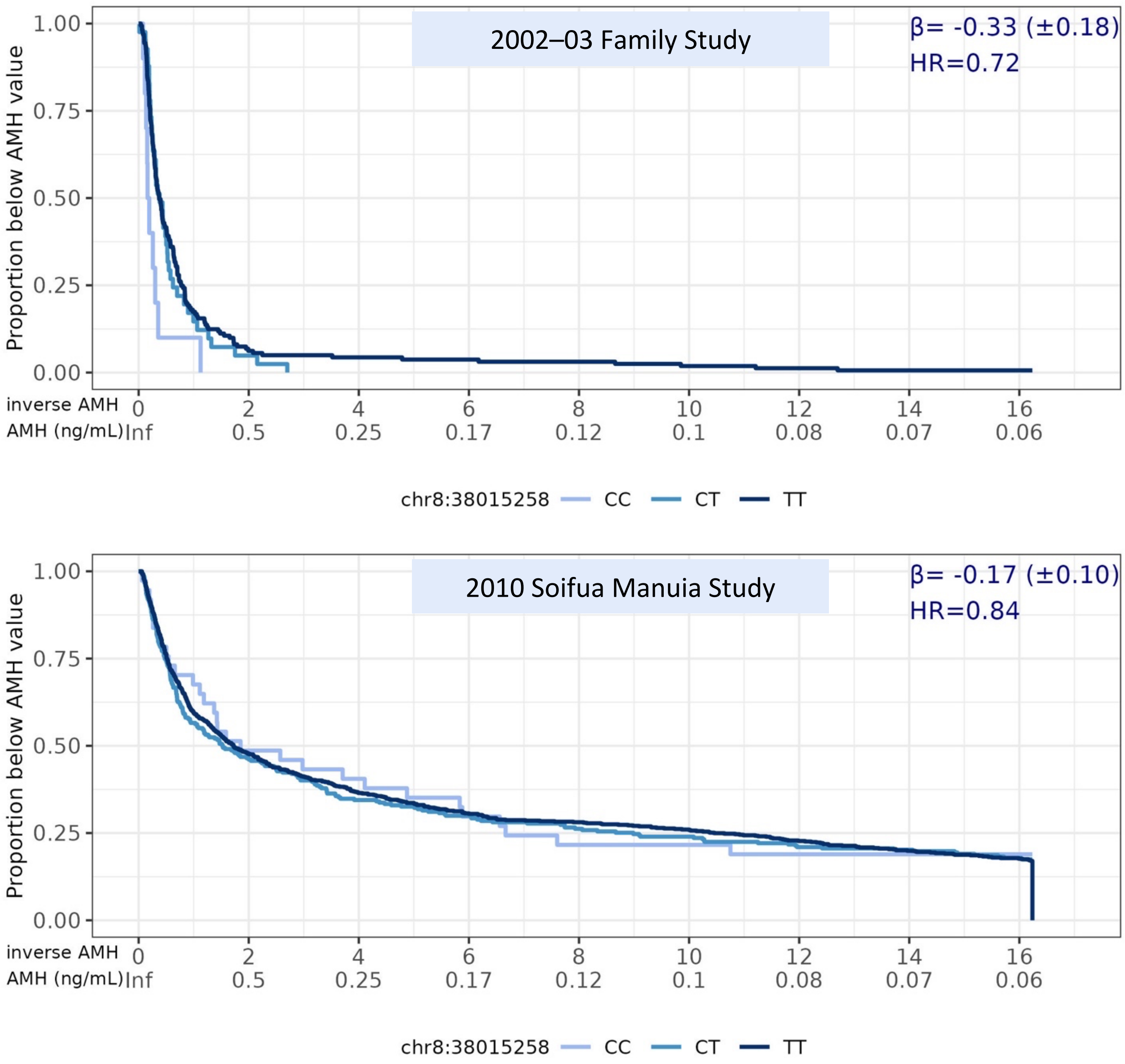

**Supplementary Figure S8 AMH levels by genotype strata for 2002–03 Family Study (top) and 2010 Soifua Manuia Study (bottom) using survival curve**

The inverse (or reciprocal) of the AMH level is shown in the *x* axis with the corresponding AMH levels in ng/mL beneath.

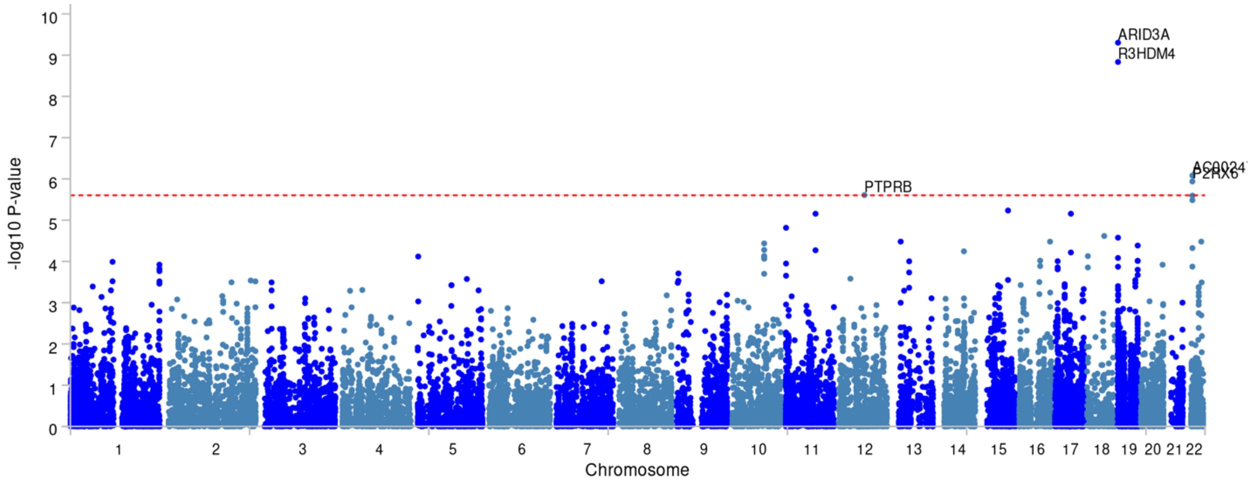

**Supplementary Figure S9 Manhattan plot of the gene-based test of meta-analysis summary statistics via MAGMA implemented in FUMA**

**Supplementary Table S2 GWAS results with *p* values < 1 × 10⁻⁵**

| Locus Information | | | | | | | 2002–03 Family Study | | | 2010 Soifua Manuia Study | | | Meta-Analysis |
|---|---|---|---|---|---|---|---|---|---|---|---|---|---|
| Variant (hg38) | Nearest Gene | Type | RDB | EA | EAF | EUR EAF | β (SE) | *p* | $R^2$ | β (SE) | *p* | $R^2$ | *p* |
| Lead SNVs from GWAS in 2002–03 Family Study | | | | | | | | | | | | | |
| 1-230483351-A-G | *PGBD5* *COG2* | intergenic | 6 | A | 0.945 | 0.940 | −1.12 (0.24) | $1.79 \times 10^{-6}$ | 0.99 | −0.1 (0.19) | $6.05 \times 10^{-1}$ | 0.98 | $1.28 \times 10^{-2}$ |
| 5-163318201-G-C | *CCNG1* | intergenic | 6 | C | 0.751 | 0.463 | −0.73 (0.16) | $3.70 \times 10^{-6}$ | 0.95 | −0.02 (0.08) | $7.84 \times 10^{-1}$ | 0.96 | $2.74 \times 10^{-2}$ |
| 6-16119809-G-C | *MYLIP* | intergenic | 7 | C | 0.132 | 0.079 | 0.9 (0.19) | $1.56 \times 10^{-6}$ | 0.99 | 0.09 (0.11) | $4.48 \times 10^{-1}$ | 0.99 | $6.53 \times 10^{-3}$ |
| 6-123454047-G-A | *TRDN* | intronic | 7 | A | 0.056 | 0.301 | 1.28 (0.29) | $6.31 \times 10^{-6}$ | 0.99 | 0.04 (0.17) | $7.98 \times 10^{-1}$ | 1.00 | $3.22 \times 10^{-2}$ |
| 6-152961137-TG-T | *FBXO5* | intergenic | | - | 0.050 | 0.000 | 1.33 (0.28) | $1.15 \times 10^{-6}$ | 0.99 | 0.18 (0.19) | $3.42 \times 10^{-1}$ | 1.00 | $3.52 \times 10^{-3}$ |
| 7-117114634-G-A | *ST7* | intronic | 5 | A | 0.091 | 0.266 | 1.01 (0.23) | $7.92 \times 10^{-6}$ | 0.96 | 0.05 (0.12) | $7.03 \times 10^{-1}$ | GT | $2.54 \times 10^{-2}$ |
| 8-5100412-C-G | *CSMD1* | intergenic | 7 | C | 0.860 | 0.854 | −1.04 (0.19) | $4.10 \times 10^{-8}$ | 0.99 | 0.13 (0.1) | $1.97 \times 10^{-1}$ | 1.00 | $2.49 \times 10^{-1}$ |
| 8-132702524-G-A | *TMEM71* | intergenic | 6 | A | 0.585 | 0.345 | −0.57 (0.13) | $6.62 \times 10^{-6}$ | 1.00 | −0.03 (0.08) | $7.30 \times 10^{-1}$ | 1.00 | $2.65 \times 10^{-2}$ |
| 9-87401964-G-A | *DAPK1* | intergenic | 5 | A | 0.064 | 0.226 | 1.28 (0.28) | $2.06 \times 10^{-6}$ | 0.99 | 0.02 (0.14) | $8.65 \times 10^{-1}$ | 1.00 | $3.06 \times 10^{-2}$ |
| 10-117766903-A-G | *EMX2* *CASC2* | intergenic | 7 | A | 0.828 | 0.860 | −0.79 (0.18) | $6.29 \times 10^{-6}$ | 0.98 | 0.02 (0.1) | $8.77 \times 10^{-1}$ | 0.99 | $7.67 \times 10^{-2}$ |
| 11-12186846-A-G | *MICAL2* | intronic | 7 | A | 0.749 | 0.796 | 0.70 (0.16) | $8.16 \times 10^{-6}$ | GT | 0.06 (0.09) | $4.87 \times 10^{-1}$ | 0.98 | $1.18 \times 10^{-2}$ |
| 11-67922682-C-T | *UNC93B1* *ALDH3B1* | intergenic | 6 | T | 0.052 | 0.000 | 1.19 (0.28) | $8.43 \times 10^{-6}$ | 0.99 | 0.28 (0.16) | $9.36 \times 10^{-2}$ | 0.95 | $6.66 \times 10^{-4}$ |
| 11-74857380-CA-C | *XRRA1* | intronic | | - | 0.939 | 0.853 | −1.24 (0.28) | $7.58 \times 10^{-6}$ | 0.92 | −0.02 (0.17) | $9.15 \times 10^{-1}$ | 0.94 | $4.66 \times 10^{-2}$ |
| 11-131774039-G-T | *NTM* | intronic | 6 | T | 0.507 | 0.502 | −0.65 (0.13) | $3.90 \times 10^{-7}$ | 0.97 | −0.05 (0.08) | $4.73 \times 10^{-1}$ | 1.00 | $5.16 \times 10^{-3}$ |
| 13-48149322-A-G | *MED4* *RB1* | intergenic | 6 | A | 0.925 | 0.955 | −1.10 (0.26) | $9.61 \times 10^{-6}$ | 0.99 | −0.02 (0.14) | $8.96 \times 10^{-1}$ | 0.99 | $4.65 \times 10^{-2}$ |
| 13-62687736-G-C | *LINC00448* | ncRNA intronic | 6 | C | 0.314 | 0.298 | −0.69 (0.14) | $8.60 \times 10^{-7}$ | 0.99 | −0.08 (0.08) | $3.05 \times 10^{-1}$ | 1.00 | $2.60 \times 10^{-3}$ |
| 14-77338499-C-T | *TMED8* | down-stream | 5 | T | 0.948 | 0.819 | −1.71 (0.29) | $3.65 \times 10^{-10}$ | 0.98 | 0 (0.18) | 1.00 | 0.99 | $8.01 \times 10^{-3}$ |
| 15-73633622-A-G | *NPTN* | upstream | 4 | A | 0.948 | 1.000 | −1.32 (0.29) | $1.83 \times 10^{-6}$ | 0.92 | 0.12 (0.19) | $5.35 \times 10^{-1}$ | 0.95 | $1.45 \times 10^{-1}$ |
| 16-50822152-C-T | *CYLD* | intergenic | 7 | T | 0.207 | 0.828 | −0.79 (0.18) | $7.75 \times 10^{-6}$ | 1.00 | 0.05 (0.09) | $5.59 \times 10^{-1}$ | 0.99 | $1.73 \times 10^{-1}$ |
| 16-57904931-C-CT | *CNGB1* | intronic | | T | 0.085 | 0.003 | 1.11 (0.23) | $1.11 \times 10^{-6}$ | 0.94 | 0.23 (0.15) | $1.24 \times 10^{-1}$ | 0.93 | $5.55 \times 10^{-4}$ |
| 19-6601983-C-T | *CD70* | intergenic | 5 | T | 0.756 | 0.699 | 0.69 (0.15) | $1.97 \times 10^{-6}$ | 0.98 | 0.03 (0.09) | $7.03 \times 10^{-1}$ | 0.98 | $1.84 \times 10^{-2}$ |
| X-2162591-A-G | *DHRSX* | intergenic | | A | 0.941 | 0.994 | −1.28 (0.29) | $5.96 \times 10^{-6}$ | 0.79 | −0.02 (0.17) | $9.14 \times 10^{-1}$ | 0.76 | $4.41 \times 10^{-2}$ |
| X-151747651-C-T | *CNGA2* | down-stream | | T | 0.374 | 0.343 | −0.64 (0.14) | $7.50 \times 10^{-6}$ | GT | −0.1 (0.08) | $1.91 \times 10^{-1}$ | 1.00 | $2.07 \times 10^{-3}$ |

| Locus Information | | | | | | | 2002–03 Family Study | | | 2010 Soifua Manuia Study | | | Meta-Analysis |
|---|---|---|---|---|---|---|---|---|---|---|---|---|---|
| Lead SNVs from GWAS in 2010 Soifua Manuia Study | | | | | | | | | | | | | |
| Variant (hg38) | Nearest Gene | Type | RDB | EA | EAF | EUR EAF | β (SE) | *p* | $R^2$ | β (SE) | *p* | $R^2$ | *p* |
| 2-36088948-A-G | *CRIM1* | intergenic | 5 | A | 0.974 | * | | | | 1.18 (0.27) | $7.15 \times 10^{-6}$ | 0.99 | |
| 2-174701241-G-A | *(GPR155) WIPF1* | intergenic | 7 | A | 0.807 | 0.913 | 0.00 (0.17) | $9.89 \times 10^{-1}$ | 0.98 | 0.45 (0.09) | $1.93 \times 10^{-6}$ | 0.99 | $1.56 \times 10^{-5}$ |
| 2-241065154-C-T | *SNED1* | intronic | 5 | T | 0.288 | 0.003 | −0.10 (0.15) | $5.30 \times 10^{-1}$ | 0.99 | −0.38 (0.08) | $7.13 \times 10^{-6}$ | 0.99 | $1.46 \times 10^{-5}$ |
| 3-19972577-A-G | *RAB5A* | intronic | 5 | A | 0.838 | * | 0.09 (0.16) | $5.86 \times 10^{-1}$ | 0.98 | −0.49 (0.1) | $1.05 \times 10^{-6}$ | 0.98 | $2.74 \times 10^{-5}$ |
| 3-190080320-A-G | *TP63 P3H2* | intronic | 7 | A | 0.910 | 0.804 | −0.27 (0.21) | $1.90 \times 10^{-1}$ | 0.99 | 0.62 (0.14) | $7.84 \times 10^{-6}$ | GT | $4.73 \times 10^{-4}$ |
| 4-18803704-C-CA | *LCORL SLIT2* | intergenic | | CA | 0.477 | 0.713 | −0.10 (0.13) | $4.18 \times 10^{-1}$ | 0.99 | 0.35 (0.08) | $3.27 \times 10^{-6}$ | 0.99 | $1.07 \times 10^{-4}$ |
| 4-41798742-G-A | *LIMCH1 PHOX2B TMEM33* | intergenic | 7 | A | 0.667 | 0.599 | −0.03 (0.14) | $8.35 \times 10^{-1}$ | GT | 0.37 (0.08) | $6.20 \times 10^{-6}$ | 0.89 | $6.15 \times 10^{-5}$ |
| 4-188298237-A-G | *TRIML1* | intergenic | 5 | A | 0.972 | 0.812 | | | | 1.10 (0.24) | $4.64 \times 10^{-6}$ | 1.00 | |
| 6-5667273-C-A | *FARS2* | intronic | 7 | A | 0.990 | 0.963 | | | | −1.74 (0.38) | $2.27 \times 10^{-6}$ | 0.92 | |
| 6-58229055-C-G | *(PRIM2) NONE* | intergenic | 7 | C | 0.967 | 0.732 | | | | 0.97 (0.22) | $8.84 \times 10^{-6}$ | 0.99 | |
| 8-14521938-T-C | *SGCZ* | intronic | 7 | T | 0.969 | * | | | | −1.01 (0.22) | $2.41 \times 10^{-6}$ | 0.98 | |
| 9-101892530-C-T | *GRIN3A* | intergenic | 7 | T | 0.024 | * | | | | 1.13 (0.23) | $7.29 \times 10^{-7}$ | 0.99 | |
| 9-107597705-C-A | *KLF4* | intergenic | 5 | A | 0.037 | * | | | | 0.89 (0.19) | $4.30 \times 10^{-6}$ | 0.97 | |
| 13-38915488-C-T | *FREM2* | intergenic | 4 | T | 0.044 | 0.022 | | | | 0.90 (0.18) | $6.66 \times 10^{-7}$ | 1.00 | |
| 13-72599097-A-ATG | *SNORA9 MZT1* | intergenic | | A | 0.634 | 1.000 | 0.20 (0.14) | $1.47 \times 10^{-1}$ | 0.96 | −0.36 (0.08) | $8.41 \times 10^{-6}$ | 0.97 | $6.19 \times 10^{-4}$ |
| 13-105438871-GAA-G | *DAOA-AS1* | intergenic | | G | 0.930 | 0.672 | 0.24 (0.27) | $3.64 \times 10^{-1}$ | 0.95 | −0.68 (0.14) | $5.75 \times 10^{-7}$ | 0.98 | $3.38 \times 10^{-5}$ |
| 14-75199954-T-C | *TMED10 FOS* | intergenic | 7 | T | 0.864 | 0.454 | 0.04 (0.19) | $8.32 \times 10^{-1}$ | 0.90 | −0.49 (0.11) | $8.01 \times 10^{-6}$ | 0.98 | $7.61 \times 10^{-5}$ |
| 15-55158410-C-T | *RSL24D1* | intergenic | 6 | T | 0.293 | 0.024 | 0.21 (0.14) | $1.40 \times 10^{-1}$ | 0.98 | −0.37 (0.08) | $3.58 \times 10^{-6}$ | 1.00 | $3.50 \times 10^{-4}$ |
| 15-97854929-G-A | *LINC00923* | ncRNA intronic | 7 | A | 0.063 | 0.000 | 0.14 (0.29) | $6.31 \times 10^{-1}$ | 0.95 | −0.72 (0.15) | $1.69 \times 10^{-6}$ | 0.96 | $3.54 \times 10^{-5}$ |
| 16-85356782-G-A | *CIBAR2 GSE1* | intergenic | 7 | A | 0.102 | 0.458 | 0.11 (0.2) | $5.88 \times 10^{-1}$ | 0.99 | 0.60 (0.13) | $2.92 \times 10^{-6}$ | 0.98 | $7.95 \times 10^{-6}$ |
| 18-77113272-G-A | *MBP* | intronic | 7 | A | 0.175 | 0.419 | −0.06 (0.18) | $7.43 \times 10^{-1}$ | 0.99 | 0.41 (0.09) | $8.44 \times 10^{-6}$ | 0.99 | $9.72 \times 10^{-5}$ |

Each SNV is presented with its gnomAD ID. The effect allele (**EA**) and its frequency in Samoans (**EAF**) followed by its frequency in Europeans in the 1000 Genome (**EUR EAF**) are reported. The effect estimates (**β**) and standard errors (**SE**) are presented except where variant MAFs were below required thresholds. **RBD** represents a score for SNV's functionality from RegulomeDB. The imputation quality score (***$R^2$***) is provided for imputed SNVs, where **GT** refers to genotyped variants

### Supplementary Table S3 Look-up of known AMH loci in Samoan GWAS.

| AMH loci from previous GWASs of European ancestry | | | | | | | | Samoan GWAS | | | | Most significant variant within ±50 kb of the AMH loci identified in previous GWASs | | | | | | | | Samoan GWAS | | |
|---|---|---|---|---|---|---|---|---|---|---|---|---|---|---|---|---|---|---|---|---|---|---|
| rsID | gnomAD ID (hg38) | Nearest Gene | EA | EUR EAF | β (SE) | *p* value | Study | $R^2$ | Samoan EAF | Effect Direction | *P* | gnomAD ID (hg38) | Functional Conse-quence | Distance (bp) | EA | Samoan EAF | Samoan LD ($r^2$) | EUR EAF gnomAD | EUR LD ($r^2$) | Effect Direction | *P* | Bonferroni corrected threshold |
| rs6729614 | 2-144887307-A-G | *TEX41* | G | 0.26 | 0.08 (0.01) | $5.56 \times 10^{-11}$ | 1 | 1.00 | 0.012 | Not examined | | 2-144839456-A-T | ncRNA intronic | −47,851 | A | 0.81 | 0.015 | 0.3783 | 0.0874 | −− | 0.0018 | 0.0028 |
| rs11683493 | 2-173394597-C-T | *CDCA7* | T | 0.57 | −0.08 (0.01) | $1.68 \times 10^{-8}$ | 2 | 1.00 | 0.349 | −+ | 0.573 | | | | | | | | | | | |
| rs116090962 | 5-146560687-G-A | *CTB-99A3.1* | A | 0.02 | 0.38 (0.07) | $6.00 \times 10^{-9}$ | 2 | 0.99 | 0.0003 | Not examined | | no variant with $p < 0.05$ within ±50 kb of rs116090962 | | | | | | | | | | |
| **rs10093345** | **8-38015258-C-T** | ***EIF4EBP1*** | **T** | **0.72** | **−0.08 (0.01)** | $\mathbf{5.05 \times 10^{-9}}$ | 1 | **0.97** | **0.825** | **−−** | **0.016** | | | | | | | | | | | |
| rs762643 | 14-53956049-G-T | *BMP4* | T | 0.44 | −0.07 (0.01) | $3.99 \times 10^{-9}$ | 1 | 1.00 | 0.338 | +− | 0.499 | | | | | | | | | | | |
| rs10417628 | 19-2251818-T-C | *AMH* | C | 0.97 | 0.32 (0.04) | $9.56 \times 10^{-12}$ | 1 | 0.09 | 0.996 | Not examined | | 19-2250470-G-A | intronic | −1,348 | A | 0.47 | 0.004 | 0.0419 | 0.0009 | ++ | 0.0006 | 0.0011 |
| rs16991615 | 20-5967581-G-A | *MCM8* | A | 0.06 | 0.16 (0.02) | $4.68 \times 10^{-9}$ | 1 | 1.00 | 0.0007 | Not examined | | 20-5993582-A-G | p.R773G | 26,001 | A | 0.83 | 0.001 | 0.00001 | * | ++ | 0.0299 | 0.0026 |
| rs186430430 | 22-28707610-T-C | *CHEK2* | C | 0.002 | 0.79 (0.12) | $9.69 \times 10^{-11}$ | 1 | 0.08 | 0.00002 | Not examined | | 22-28703766-T-C | intronic | −3,844 | T | 0.84 | NA | 0.4825 | 0.0037 | −− | 0.0051 | 0.0019 |

The variant info for the lead SNVs identified in AMH GWASs from women of European ancestry (**EUR**) is under the light blue header. For the lead variant in each known AMH locus, we report the **rsID**, **gnomAD ID**, **nearest gene**, effect allele (**EA**), effect allele frequency (**EAF**) in the originating study, effect size and standard error (**β (SE)**), ***p* value**, and reference to the originating study: [1)] (Pujol-Gualdo *et al.*, 2024) [2)] (Verdiesen *et al.*, 2022)

Next, under the center left dark blue header, we provide imputation quality (***R*** $^{2}$), **EAF** in Samoans, **effect directions** in the meta-analyzed samples, and the ***p* value** from the **Samoan GWAS** for the same SNV, if the allele frequency of the SNV was high enough to be included in the meta-analysis (see Methods).

The next seven columns contain information about the lead variant in the Samoan GWAS within ±50 kb of the lead variant from prior European GWASs if the lead variant was not examined in the Samoan GWAS and if the nearby variant had $p < 0.05$. For each SNV, we provide the **gnomAD ID**, **functional consequence**, **distance** from the prior GWAS lead SNV, the **EAF** and the **LD** $r^2$ with the prior GWAS lead SNV in Samoans, and the **EAF** (gnomAD v4) and **LD** $r^2$ with the prior GWAS lead SNV in Europeans (1000G EUR, calculated using LDpair function in LDlink from NIH). An asterisk (*****) indicates that the variant was not present in 1000 Genomes.

The final three columns, under the righthand dark blue header, contain the **effect directions** in the meta-analyzed samples, the ***p* value** from the Samoan GWAS, and the per-region **Bonferroni-corrected threshold** for statistical significance.

The AMH loci from previous GWASs that replicated in Samoan meta-analysis are in bold.

**Supplementary Table S4 Results from transcriptome-wide analysis**

| Gene | Chr | Top Tissue | $p$ | $z$ score | | | |
|---|---|---|---|---|---|---|---|
| | | | | Mean | SD | Min | Max |
| ***GINS2*** | **16** | **hypothalamus** | $\mathbf{2.20 \times 10^{-18}}$ | **−0.372** | **1.797** | **−1.926** | **3.125** |
| ***SENP3*** | **17** | **pituitary** | $\mathbf{2.24 \times 10^{-7}}$ | **0.382** | **2.227** | **−2.978** | **2.344** |
| ***USP7*** | **16** | **subcutaneous adipose** | $\mathbf{3.63 \times 10^{-7}}$ | **0.657** | **2.168** | **−1.805** | **2.282** |
| ***TUSC3*** | **8** | **liver** | $\mathbf{4.38 \times 10^{-7}}$ | **−0.206** | **1.287** | **−1.661** | **2.052** |
| ***MAFA*** | **8** | **subcutaneous adipose** | $\mathbf{9.30 \times 10^{-7}}$ | **−0.097** | **1.425** | **−2.141** | **1.080** |
| ***METTL4*** | **18** | **adrenal gland** | $\mathbf{9.31 \times 10^{-7}}$ | **−0.022** | **1.355** | **−2.861** | **1.191** |
| ***NDFIP1*** | **5** | **pancreas** | $\mathbf{2.34 \times 10^{-6}}$ | **0.015** | **1.798** | **−3.650** | **2.175** |
| *ACTR2* | 2 | ovary | $6.20 \times 10^{-6}$ | −0.884 | 1.437 | −1.753 | 2.096 |
| *RP11-434E6.4* | 16 | pancreas | $6.98 \times 10^{-6}$ | 1.151 | 1.496 | −2.049 | 2.875 |
| *CHMP1A* | 16 | liver | $1.17 \times 10^{-5}$ | −1.674 | 1.357 | −3.978 | −0.345 |
| *OSMR* | 5 | liver | $1.26 \times 10^{-5}$ | −0.417 | 1.618 | −2.863 | 2.632 |
| *ARID3A* | 19 | pancreas | $1.32 \times 10^{-5}$ | 3.003 | 1.168 | 1.596 | 4.494 |
| *MRPL44* | 2 | ovary | $1.44 \times 10^{-5}$ | 0.850 | 1.246 | −0.456 | 3.514 |
| *PAK4* | 19 | pancreas | $2.17 \times 10^{-5}$ | −0.076 | 1.056 | −1.590 | 0.931 |
| *RP11-329B9.4* | 3 | hypothalamus | $2.26 \times 10^{-5}$ | −0.543 | 1.311 | −2.541 | 0.806 |
| *L3MBTL1* | 20 | thyroid | $2.29 \times 10^{-5}$ | −0.386 | 1.841 | −2.855 | 2.368 |
| *HMGCS2* | 1 | pituitary | $2.46 \times 10^{-5}$ | −0.494 | 1.106 | −2.050 | 1.028 |
| *RP11-329B9.5* | 3 | pituitary | $2.72 \times 10^{-5}$ | −0.170 | 1.399 | −1.920 | 1.392 |
| *ZNF362* | 1 | subcutaneous adipose | $3.07 \times 10^{-5}$ | 0.017 | 1.369 | −2.075 | 1.193 |
| *MLH3* | 14 | adrenal gland | $3.60 \times 10^{-5}$ | 3.481 | 1.005 | 1.292 | 4.005 |
| *ABCD4* | 14 | hypothalamus | $4.12 \times 10^{-5}$ | 1.880 | 1.709 | −1.074 | 3.568 |
| *LYZ* | 12 | liver | $4.23 \times 10^{-5}$ | 0.179 | 1.197 | −1.004 | 1.776 |
| *RPP25* | 15 | subcutaneous adipose | $4.63 \times 10^{-5}$ | −0.503 | 1.023 | −1.250 | 0.637 |
| *RMI2* | 16 | ovary | $4.89 \times 10^{-5}$ | 1.925 | 1.524 | −1.037 | 2.948 |

Statistically significant findings are represented by *p* values depicted in bold.
